# Outbreaks of Covid-19 Variants in Prisons: A Mathematical Modeling Analysis of Vaccination and Re-Opening Policies

**DOI:** 10.1101/2021.05.03.21256525

**Authors:** Theresa Ryckman, Elizabeth T. Chin, Lea Prince, David Leidner, Elizabeth Long, David M. Studdert, Joshua A. Salomon, Fernando Alarid-Escudero, Jason R. Andrews, Jeremy D. Goldhaber-Fiebert

## Abstract

**Background:** Residents of correctional facilities have experienced disproportionately higher rates of SARS-CoV-2 infection and Covid-19-related mortality. To protect against outbreaks, many prisons and jails imposed heavy restrictions on in-person activities, which are now beginning to lift. Uncertainty surrounds the safety of these moves.

**Methods and Findings:** We obtained system-wide resident-day level data for the California state prison system, the nation’s third largest. We used the data to develop a transmission-dynamic stochastic microsimulation model that projects the impact of various policy scenarios on risks of SARS-CoV-2 infections and related hospitalization among residents after an initial infection is introduced to a prison. The policy scenarios vary according to levels of vaccine coverage, baseline immunity, resumption of activities, and use of non-pharmaceutical interventions (e.g., masking, physical distancing). The analyses were conducted across 5 types of prisons that differed in their residential layouts, security levels, and resident demographics.

If a viral variant is introduced into a prison that has resumed pre-2020 contact levels, has moderate vaccine coverage, and has no baseline immunity, 23-74% of residents are expected to be infected over 200 days. High vaccination coverage coupled with use of non-pharmaceutical measures reduces cumulative infections to 2-54% of residents. In prisons consisting mostly of dormitory housing, even with high vaccine coverage and non-pharmaceutical interventions, resumption of in-person activities is associated with substantial risk, unless there is high baseline immunity (e.g., ≥50%) from prior outbreaks. In prisons consisting mostly of cell housing, <10% of residents are expected to become infected, even with no baseline immunity. However, hospitalization risks are substantial in prisons that house medically vulnerable populations, even for prisons consisting mostly of cells. Risks of large outbreaks are substantially higher if there is continued introduction of infections into a prison. Some findings may not be transportable to other carceral settings, and our assumptions regarding viral variants will not be accurate for all variants.

**Conclusions:** Balancing the benefits of resuming normal in-person activities against the risks of Covid-19 outbreaks is a difficult challenge for correctional systems. The policy choices are not strictly binary. To protect against viral variants, prisons should focus on achieving both high vaccine coverage and maintaining widespread use of non-pharmaceutical interventions. With both in place, some prisons, especially those with lower room occupancy that have already had large outbreaks, could safely resume in-person activities, while continuing testing and measures to protect the medically-vulnerable.

## Introduction

In the Covid-19 pandemic’s first year, US prison populations had infection and mortality rates 5-6 and 2-3 times higher, respectively, than free-living populations (1–3). Overcrowded congregate living spaces, inadequate testing, lack of personal protective equipment and adequate sanitation, mistrust of medical personnel, and policies that disincentivize symptom reporting increase outbreak risks in prisons (3,4). Non-pharmaceutical strategies (e.g. masking and distancing) to reduce transmission are less feasible in such settings, which often house populations that are more medically vulnerable than the general population. Incarcerated individuals living in large dormitories are at especially high risk for infection (5,6).

Whether prisons already have sufficient natural immunity from prior outbreaks to safely resume pre-epidemic activities without first achieving high vaccination levels is an open question (7–9). Two epidemiological features suggest otherwise.

First, the threshold level of natural immunity required to prevent subsequent outbreaks depends on contacts (10). This threshold is likely higher in prisons than in the general population given density and crowding. Since 2020, many prisons halted in-person activities, including group therapy, and outside visitation, to reduce contacts and transmission. While resumption of these activities will yield tangible benefits, it also increases contact and transmission risks that could overwhelm the protection conferred by built-up immunity.

Second, SARS-CoV-2 viral variants have emerged that are more transmissible, more likely to result in severe disease, and can evade immunity from prior infection (11–13). Genetic sequencing data indicate that more transmissible variants’ share of total Covid-19 cases in the US is growing exponentially (14,15). The paucity of genetic sequencing in most prisons (3) increases the risk that a new variant will trigger a large outbreak before correctional systems are aware of the introduction, limiting the efficacy of reactive measures like mass testing and quarantine.

Whether current vaccination levels in carceral settings are sufficient to halt transmission is thus unclear (16). As states scale-up vaccination for incarcerated populations (17,18), acceptance will determine achievable coverage. Even in states where incarcerated people have been prioritized for vaccination, hesitancy remains a concern (3,19). Our study uses simulation modeling to assess how the size and consequences of Covid-19 outbreaks stemming from the introduction of a viral variant into a prison depend on prior immunity, vaccine coverage, and whether and how activities are resumed.

## Methods

### Overview

Using a mathematical model, we assess the impact of vaccination on Covid-19 in US prisons. We examine how three factors influence risks of outbreaks and severe outcomes: (1) resumption of in-person activities; (2) use of non-pharmaceutical interventions (NPIs) such as face coverings and physical distancing; and (3) vaccination among incarcerated individuals. We also assess how two exogeneous factors - the introduction of a highly transmissible viral variant and baseline natural immunity from prior outbreaks - affect these estimates. We use detailed data from the California state prison system and facilitate generalizability to other prison systems by simulating a range of prisons that differ in room occupancies, residential layouts, resident demographics, security levels, and contact patterns.

### Model

The transmission-dynamic stochastic microsimulation follows individual residents and correctional staff within a prison (Figure 1, Appendix). We simulate different prisons by instantiating the model with prison-specific characteristics. These include the prison’s residential layout: composition of rooms (i.e., cells or dorms) and room occupancy – which has been identified as an especially important risk factor for prison outbreaks (5,6) – and their organization within buildings and yards. They also include the number of residents and their age, sex, comorbidities, security level, room assignment and type, and participation in prison labor or other activities. Correctional staff are characterized by age; the size of the staff population varies by prison. We selected prisons from California’s 35 state prisons that demonstrate variation across these characteristics and are comparable to other prison systems in the US. Simulated prisons include: a low to medium security men’s prison consisting mostly of dormitories (defined as rooms with ≥3 occupants), a low to medium security men’s prison with a mix of dorms and cells (rooms with ≤2 occupants), a high security men’s prison with mostly cells, a women’s prison with mixed security levels and mostly cells, and a medical prison that houses older residents and residents with medical vulnerabilities mostly in cells (Appendix).

**Figure 1:**
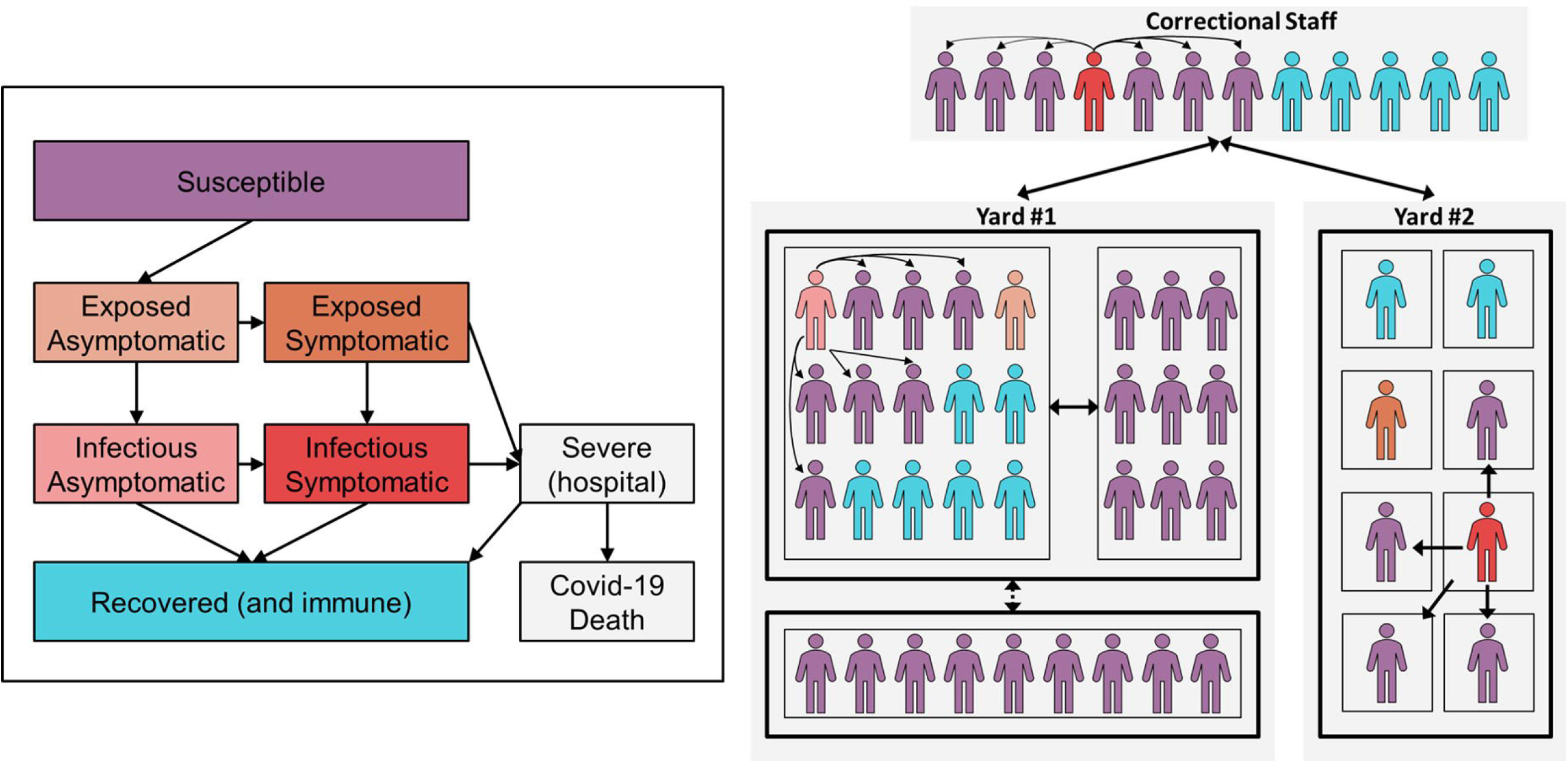
Prison microsimulation model diagram. The left panel shows possible state transitions and outcomes for an individual who becomes infected. The right panel shows the distributions and transmission of individuals within a prison. Thick black boxes on the right denote buildings within prison yards, while thin black boxes denote rooms within buildings, and arrows designate different possible routes of transmission from infectious individuals: within-room, within-building, within-yard, staff-to-staff, staff-to-resident, and resident-to-staff.

Daily transmission risk is based on the number of contacts each person has, the proportion of contacts that are infectious, and the probability of transmission by type of infectious contact (equations in Appendix). Residents have contact with their roommates, other residents in the same building, and correctional staff across the prison. Residents who participate in out-of-room activities (labor, school, or other activities) also have contact with other residents in the same yard who participate in those activities. Transmission risks are highest for in-room contacts, followed by activity contacts, and then building and staff contacts (Table 1). Because the model reflects differences in residents’ housing and activity participation, it captures variation in individuals’ exposures and thus replicates prior study findings (5) (Appendix). Correctional staff are assumed to mix homogenously with each other and with residents.

**Table 1:**
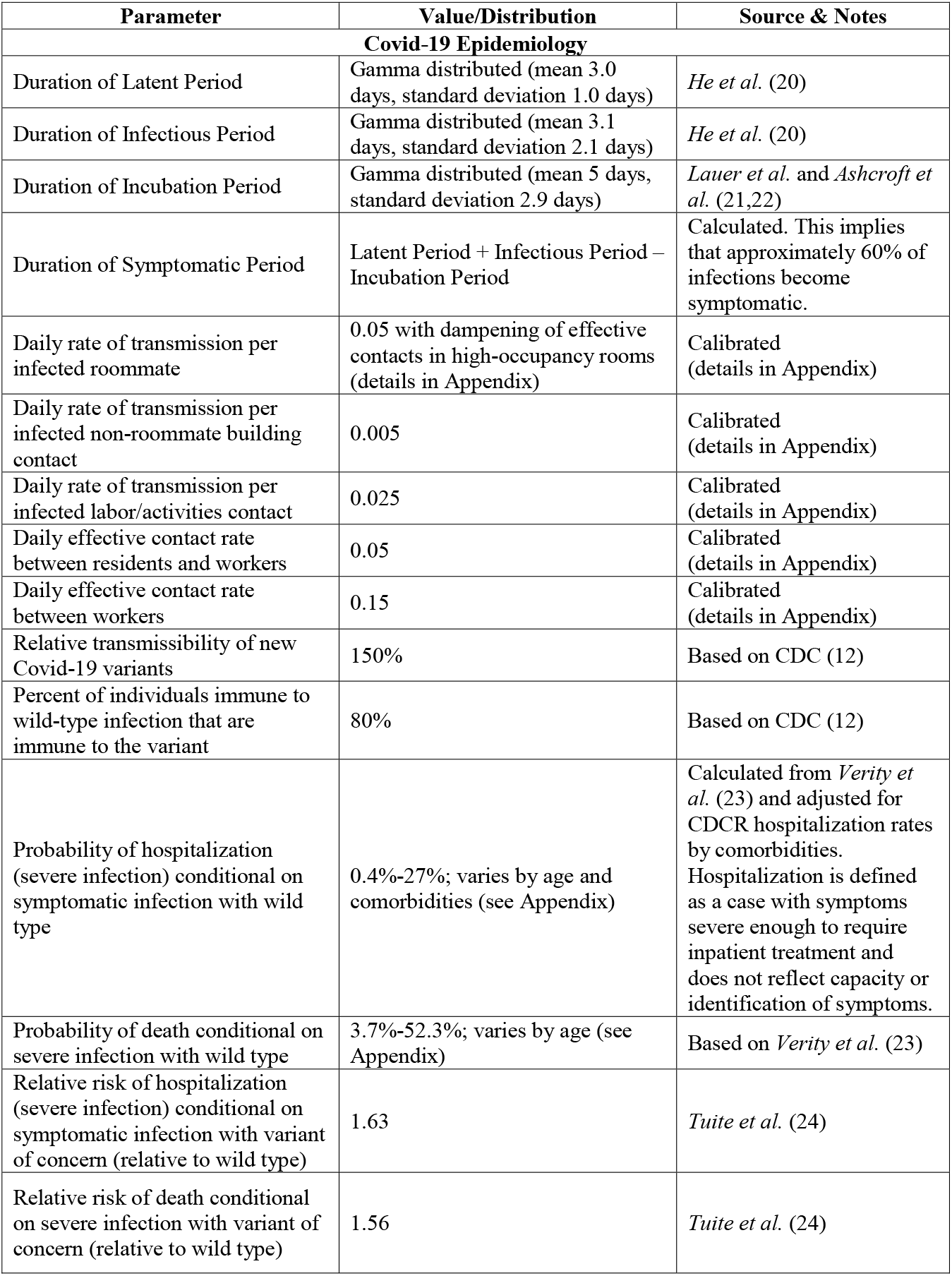
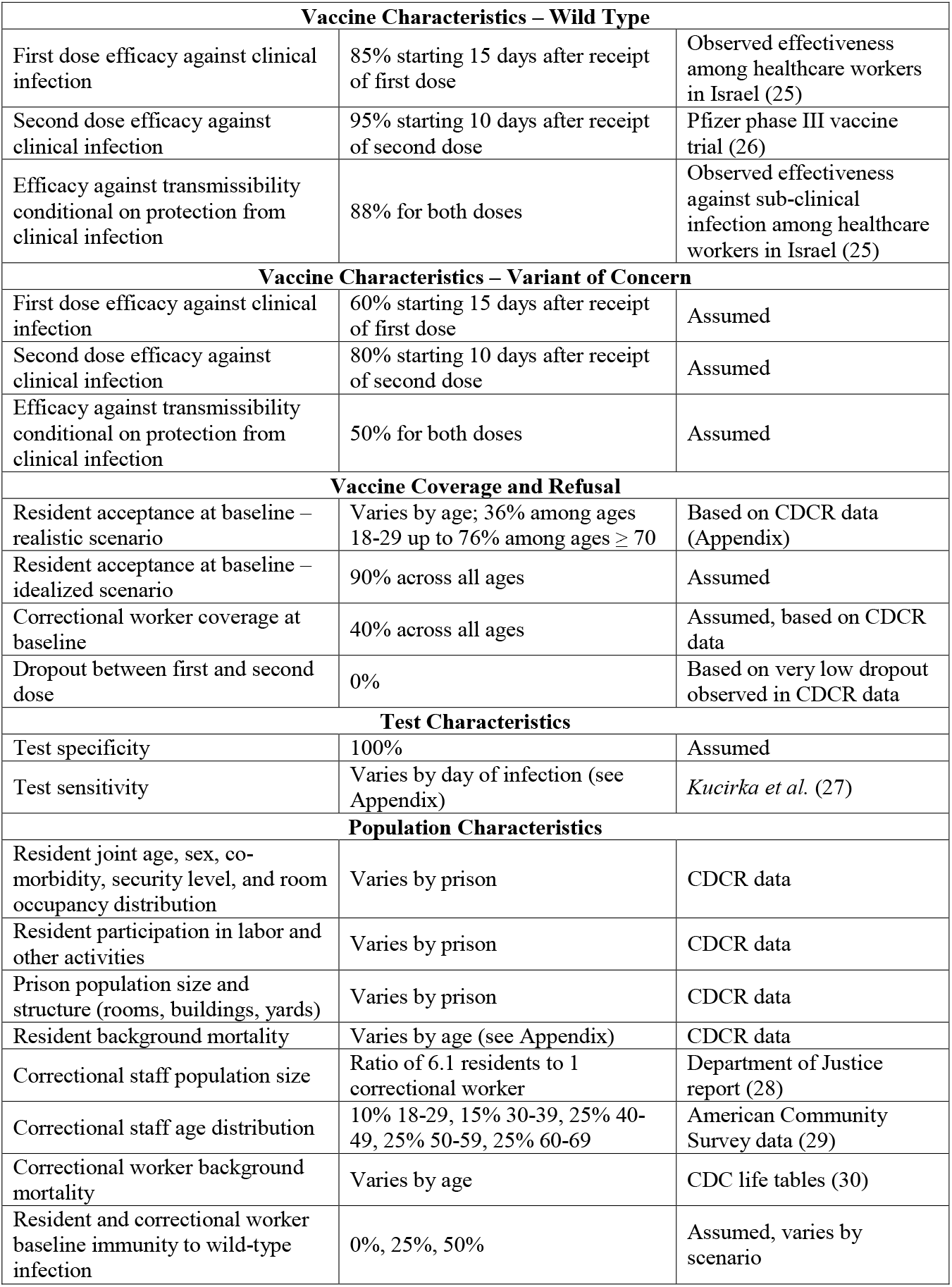
Prison microsimulation model parameters.

| Parameter | Value/Distribution | Source & Notes |
| --- | --- | --- |
| **Covid-19 Epidemiology** |  |  |
| Duration of Latent Period | Gamma distributed (mean 3.0 days, standard deviation 1.0 days) | *He et al. (20)* |
| Duration of Infectious Period | Gamma distributed (mean 3.1 days, standard deviation 2.1 days) | *He et al. (20)* |
| Duration of Incubation Period | Gamma distributed (mean 5 days, standard deviation 2.9 days) | *Lauer et al. and Ashcroft et al. (21,22)* |
| Duration of Symptomatic Period | Latent Period + Infectious Period – Incubation Period | Calculated. This implies that approximately 60% of infections become symptomatic. |
| Daily rate of transmission per infected roommate | 0.05 with dampening of effective contacts in high-occupancy rooms (details in Appendix) | Calibrated (details in Appendix) |
| Daily rate of transmission per infected non-roommate building contact | 0.005 | Calibrated (details in Appendix) |
| Daily rate of transmission per infected labor/activities contact | 0.025 | Calibrated (details in Appendix) |
| Daily effective contact rate between residents and workers | 0.05 | Calibrated (details in Appendix) |
| Daily effective contact rate between workers | 0.15 | Calibrated (details in Appendix) |
| Relative transmissibility of new Covid-19 variants | 150% | Based on CDC (12) |
| Percent of individuals immune to wild-type infection that are immune to the variant | 80% | Based on CDC (12) |
| Probability of hospitalization (severe infection) conditional on symptomatic infection with wild type | 0.4%-27%; varies by age and comorbidities (see Appendix) | Calculated from *Verity et al. (23)* and adjusted for CDCR hospitalization rates by comorbidities. Hospitalization is defined as a case with symptoms severe enough to require inpatient treatment and does not reflect capacity or identification of symptoms. |
| Probability of death conditional on severe infection with wild type | 3.7%-52.3%; varies by age (see Appendix) | Based on *Verity et al. (23)* |
| Relative risk of hospitalization (severe infection) conditional on symptomatic infection with variant of concern (relative to wild type) | 1.63 | *Tuite et al. (24)* |
| Relative risk of death conditional on severe infection with variant of concern (relative to wild type) | 1.56 | *Tuite et al. (24)* |

| **Vaccine Characteristics – Wild Type** |  |  |
| --- | --- | --- |
| First dose efficacy against clinical infection | 85% starting 15 days after receipt of first dose | Observed effectiveness among healthcare workers in Israel (25) |
| Second dose efficacy against clinical infection | 95% starting 10 days after receipt of second dose | Pfizer phase III vaccine trial (26) |
| Efficacy against transmissibility conditional on protection from clinical infection | 88% for both doses | Observed effectiveness against sub-clinical infection among healthcare workers in Israel (25) |
| **Vaccine Characteristics – Variant of Concern** |  |  |
| First dose efficacy against clinical infection | 60% starting 15 days after receipt of first dose | Assumed |
| Second dose efficacy against clinical infection | 80% starting 10 days after receipt of second dose | Assumed |
| Efficacy against transmissibility conditional on protection from clinical infection | 50% for both doses | Assumed |
| **Vaccine Coverage and Refusal** |  |  |
| Resident acceptance at baseline – realistic scenario | Varies by age; 36% among ages 18-29 up to 76% among ages $\geq 70$ | Based on CDCR data (Appendix) |
| Resident acceptance at baseline – idealized scenario | 90% across all ages | Assumed |
| Correctional worker coverage at baseline | 40% across all ages | Assumed, based on CDCR data |
| Dropout between first and second dose | 0% | Based on very low dropout observed in CDCR data |
| **Test Characteristics** |  |  |
| Test specificity | 100% | Assumed |
| Test sensitivity | Varies by day of infection (see Appendix) | *Kucirka et al. (27)* |
| **Population Characteristics** |  |  |
| Resident joint age, sex, co-morbidity, security level, and room occupancy distribution | Varies by prison | CDCR data |
| Resident participation in labor and other activities | Varies by prison | CDCR data |
| Prison population size and structure (rooms, buildings, yards) | Varies by prison | CDCR data |
| Resident background mortality | Varies by age (see Appendix) | CDCR data |
| Correctional staff population size | Ratio of 6.1 residents to 1 correctional worker | Department of Justice report (28) |
| Correctional staff age distribution | 10% 18-29, 15% 30-39, 25% 40-49, 25% 50-59, 25% 60-69 | American Community Survey data (29) |
| Correctional worker background mortality | Varies by age | CDC life tables (30) |
| Resident and correctional worker baseline immunity to wild-type infection | 0%, 25%, 50% | Assumed, varies by scenario |

### Covid-19 Epidemiology

Infected individuals become infectious over time and some develop symptoms (20–22) (Appendix). Those with symptomatic infection have age- and comorbidity-specific risks of severe Covid-19 requiring hospitalization and of Covid-19-related mortality. Individuals who survive recover and are immune to infection. Individuals also face background age- and sex-specific mortality risks (Appendix). Covid-19 natural history is the same for incarcerated individuals and correctional staff.

Reflecting California prison policies, in our model infected individuals can be detected via symptom screening, surveillance testing, or reactive testing, triggered by other cases detected nearby (Appendix). Once detected, an individual is isolated, and all other individuals residing in the building of the detected individual are placed into quarantine. Those quarantined continue to mix with each other but have no contact with other buildings. Quarantine lasts for 14 days after the last case detected in a building (Appendix). Test sensitivity varies by day since infection (27). Given the limited analytical timeframe (200 days), new admissions, releases, and transfers between prisons are not modeled, allowing the analysis to focus on the impact of a single imported infection at the start of the simulation.

### Resumption of Activities and NPIs

We model re-opening as a return to pre-Covid out-of-room activity levels and doubling of building and staff contacts, the latter based on average differences between pre-Covid and Covid activity levels. We assess re-opening with or without NPIs (e.g., facial coverings and physical distancing during in-person activities) that reduce transmission risks from contacts resulting from re-opening by 75%. These NPIs are assumed to already be in place in scenarios that keep activities at current levels.

### Covid-19 Variants and Immunity

We consider scenarios in which either a new, more transmissible and severe SARS-CoV-2 variant like *B.1.1.7* (12,13,24) or non-variant (wild type) SARS-CoV-2 infection is introduced into a prison. Scenarios are also differentiated by the size of prior outbreaks and consequent baseline immunity levels in the incarcerated and staff populations. Immunity is modeled as moderately less protective against the new variant than the wild type (Table 1).

### Vaccination

We analyze two levels of vaccine acceptance among residents (90% “idealized” acceptance, lower and age-varying “realistic” acceptance; Table 1). Based on early evidence from California prisons, we assume that 40% of staff are vaccinated and that resident and staff vaccination is achieved before an infection is imported into the prison.

Vaccine efficacy was based on *Pfizer-BioNTech* and *Moderna* trials and observational studies that indicate lower efficacy against transmissibility than against clinical infection and higher efficacy after complete vaccination, compared with efficacy after the first dose (Table 1) (25,26). We assumed the vaccine is moderately less effective against new variants than wild type Covid-19 (Table 1).

### Data

The California Department of Corrections and Rehabilitation (CDCR) provided daily individual-level data on all residents in its custody. Data describe sociodemographic characteristics, housing status, chronic conditions and health status, participation in labor and other activities, and Covid-19 testing, infection, and vaccination status. We supplemented these data with literature-based estimates related to Covid-19 epidemiology, testing, and vaccination and public data describing correctional staff (Table 1).

### Outcomes

We compared total resident infections (regardless of detection and symptoms) and severe cases (requiring hospitalization and possibly resulting in death) across scenarios and policies. We computed cumulative risks of these outcomes over 200 days.

### Technical Details

The model is constructed in R (version 3.6.3). We initiated simulations by seeding one infected resident in a prison and compared mean outcomes from 500 simulations across each scenario and prison, employing variance reduction techniques (31).

### Human Subjects Research

Stanford University’s Institutional Review Board approved the protocol (IRB-55835).

## Results

### Infections

Risks of Covid-19 outbreaks are highest when a viral variant is introduced to a prison with low baseline immunity where only moderate but realistic vaccination coverage has been achieved and activities have been reopened absent NPIs (Figure 2, panel A). In these settings, the percentage of residents infected over 200 days ranges from 23% in a medical prison with mostly cells and protected vulnerable populations, to 54-55% in celled prisons with greater activity participation than medical settings (including the women’s prison), and up to 73-74% in prisons with some or almost all dorms. Even when half of residents have immunity to wild type SARS-CoV-2 from prior infection (translating to 40% of residents protected against infection with a new variant), 25-32% of residents are expected to be infected in prisons with dorms.

**Figure 2:**
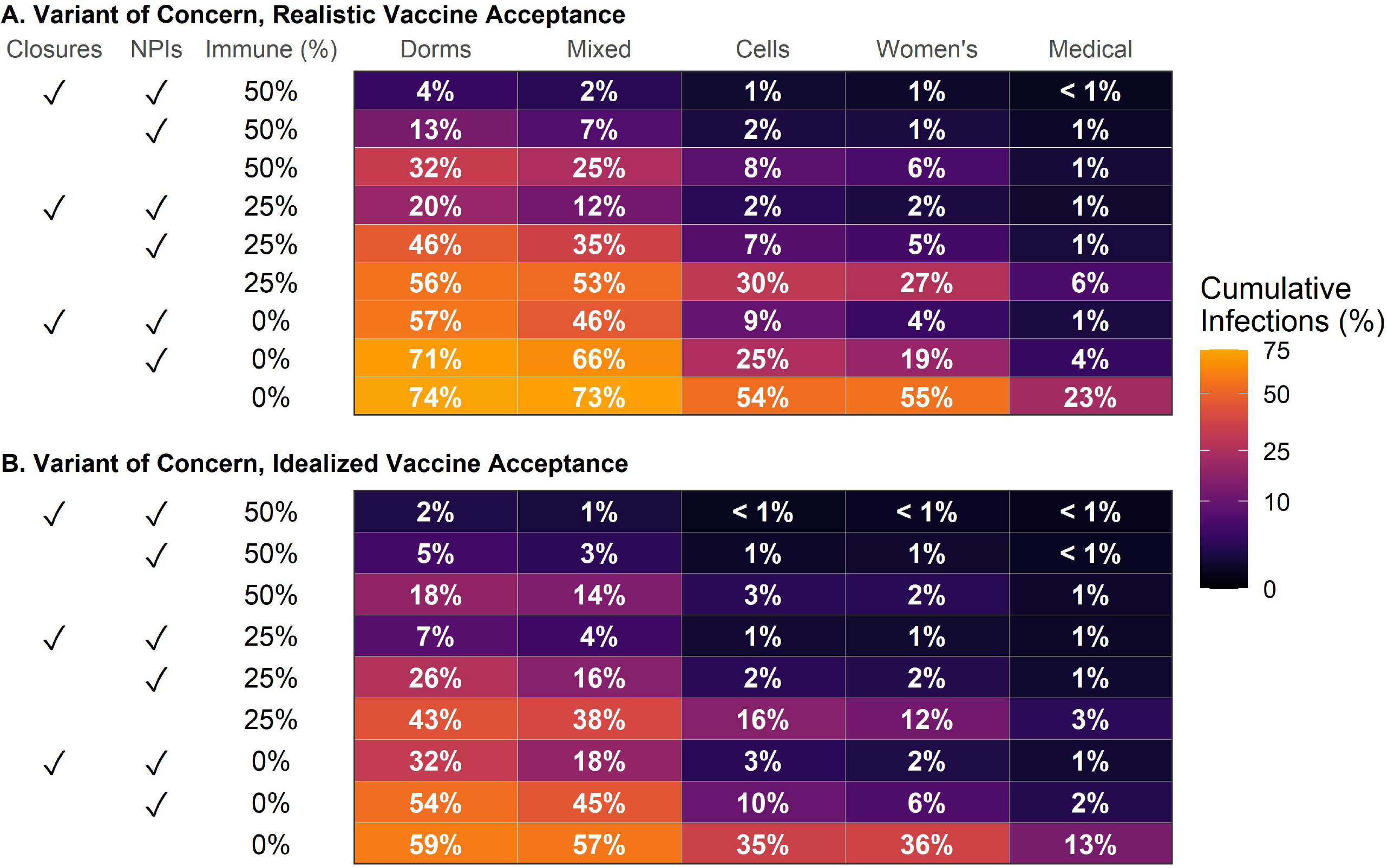
Cumulative resident infections over 200 days by in-person activity status, widespread use on NPIs, and baseline immunity, conditional on introduction of a single variant infection. Figure shows average cumulative infections among residents across 500 model simulations over 200 days for each scenario shown.

NPIs that are 75% effective in reducing contact transmissibility can reduce infections in settings with realistic vaccine coverage but alone are not sufficient to counteract risks from resumption of activities. Even when activities and other protections remain at their current levels, in prisons with low baseline immunity infection levels of 4-9% are expected in the men’s prison with mostly cells and the women’s prison (which also has mostly cells) up to 46-57% in prisons with dorms. With reopening and NPIs, these levels increase to 19-25% and 66-71%, respectively.

Achieving 90% vaccination coverage of residents substantially reduces expected outbreak sizes (Figure 2, panel B). The largest reductions from high vaccine coverage relative to realistic coverage are in settings that already have high baseline immunity and/or do not resume in-person activities. In these settings, high vaccination coverage yields an approximately two-fold reduction in infections. In settings with lower immunity that re-open, achieving high vaccine coverage is still beneficial but has less impact. Additionally, NPIs that are 75% effective are most impactful in settings where vaccination coverage and baseline immunity are already high. Thus, NPIs and vaccination are complementary interventions, not substitutes.

### Hospitalizations and Deaths

Severe outcomes follow similar patterns to infections, with several notable exceptions (Figure 3). The highest hospitalization levels are expected when a new variant is introduced to a prison with dorms and low baseline immunity that has resumed activities without NPI use (Figure 3 panel A). In these settings, with realistic vaccine acceptance, between 12 and 17 hospitalizations per 1000 residents are expected. Achieving high vaccination coverage effectively halves these hospitalizations (6-10 per 1000; Figure 3 panel B). High vaccination coverage reduces hospitalization to an even greater degree when these prisons with dorms do not resume activities (9-10 per 1000 with realistic vaccine acceptance vs. 3 per 1000 with idealized vaccine acceptance). Because vaccines yield greater protection against symptomatic infections than transmissibility, they have a larger impact on incidence of severe outcomes than on infections.

**Figure 3:**
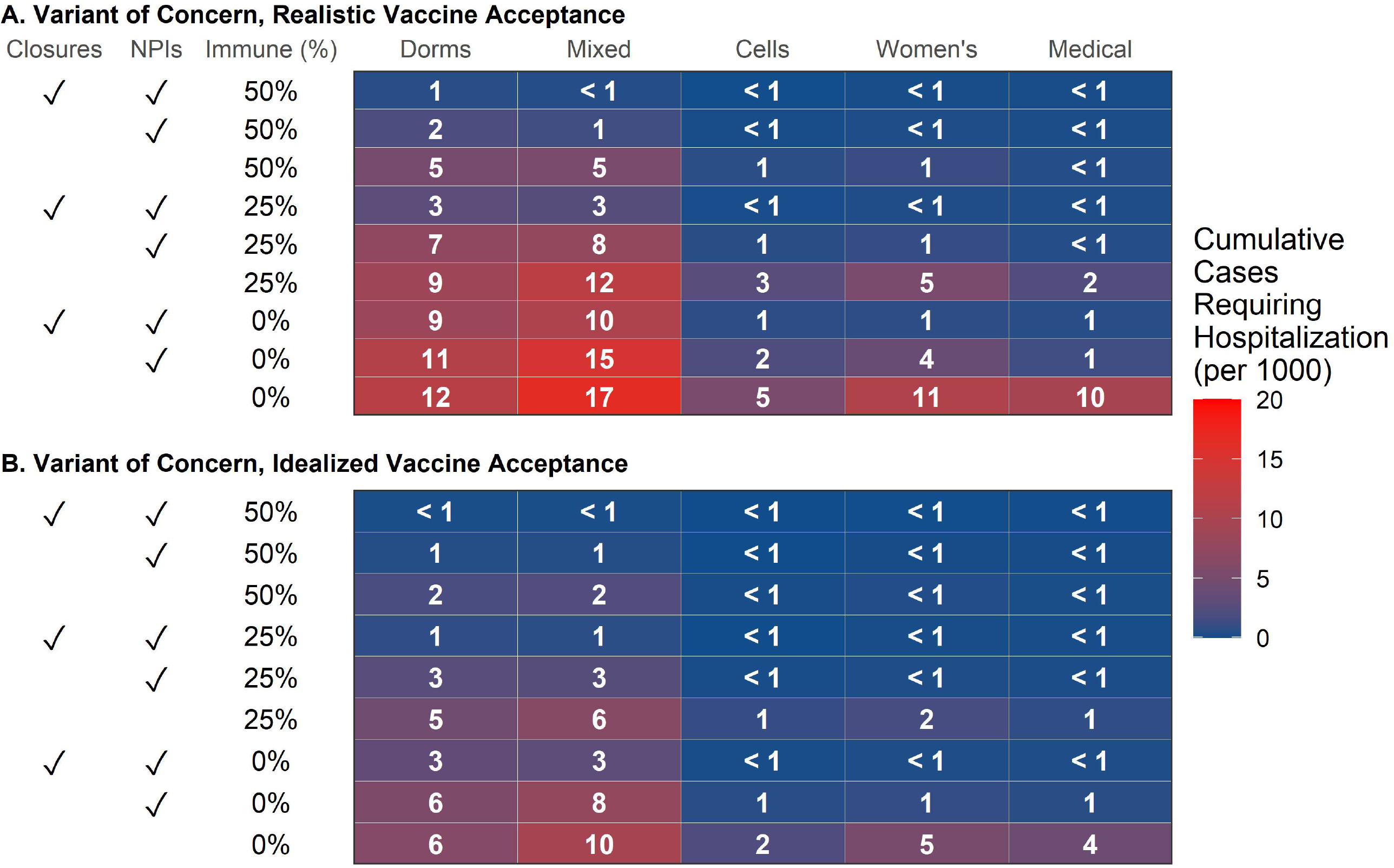
Cumulative resident cases requiring hospitalization over 200 days by in-person activity status, widespread use on NPIs, and baseline immunity, conditional on introduction of a single variant infection. Figure shows average cumulative severe cases (requiring hospitalization) among residents across 500 model simulations over 200 days for each scenario shown.

In each scenario, medical prisons are expected to have the lowest levels of cumulative infections among the five prison types. In addition to having more protections in place than other prison types, residents of the medical prison we modeled mostly live in cells within many small buildings, reducing opportunities for rapid spread across the prison. However, residents of medical prisons also tend to be older and have more comorbidities, increasing their risk of severe outcomes from infection. Those vulnerabilities manifest in disproportionately high rates of hospitalization (Appendix Figure S13). While fewer infections are expected in medical prisons, hospitalizations are in some scenarios double those in the high security prison with mostly cells. With re-opening absent NPI use, low baseline immunity, and realistic vaccination levels, hospitalizations in medical prisons could be as high as 23 per 1000 residents, translating to almost 20 Covid-related deaths in a prison of 3000 people (Appendix Figure S14).

Women’s prisons are similar to medical prisons in this respect. Since California has no women’s medical prison, medically vulnerable women are housed in one of two general population women’s prisons. While the women’s prison we model has mostly cells and similar infection levels as the high security men’s prison with mostly cells, it experiences higher rates of hospitalizations.

Vaccination reduces the proportion of infections that require hospitalization and hence, compared to no vaccination, conveys additional benefits to older and medically vulnerable residents, both because they are at greater risk of adverse outcomes and because they have observably higher levels of vaccine acceptance (Appendix Figures S13, S15-S16).

Cumulative infections and severe cases are expected to be substantially lower if a wild type infection is introduced to a prison, compared to a new variant introduction (Appendix Figures S17-S18). With more realistic vaccine coverage, prisons with low baseline immunity, especially those with dorms, are still at risk of large outbreaks (19-23% of residents infected) if activities are resumed without NPIs in place. With high vaccine coverage, even settings with low baseline immunity and high levels of contacts are anticipated to have few infections (1% of residents infected or less). However, as variants predominate globally, the likelihood that infection introductions will be new variants like *B.1.1.7* grows as well.

### Sensitivity Analysis

Many prisons have experienced repeated importations of infection, in which case outbreaks are far less likely to die out. In an analysis that allowed for repeated importation, which could arise from staff, new arrivals, or transfers, increases in resident infections from resumption of in-person activities are substantially larger than in our main results (Figure 4, Appendix Figure S19). The biggest differences occur in settings that are at lower risk in the main analysis: those with high baseline immunity and NPIs in place and celled, women’s, and medical prisons. With realistic vaccine coverage, resumption of activities in these three prison types is expected to result in 5-27% of residents infected if there is high baseline immunity and 27-68% of residents infected if there is no baseline immunity, translating to substantial expected numbers of hospitalizations (i.e., 21 per 1000 residents in medical prisons; Appendix Figure S20).

**Figure 4:**
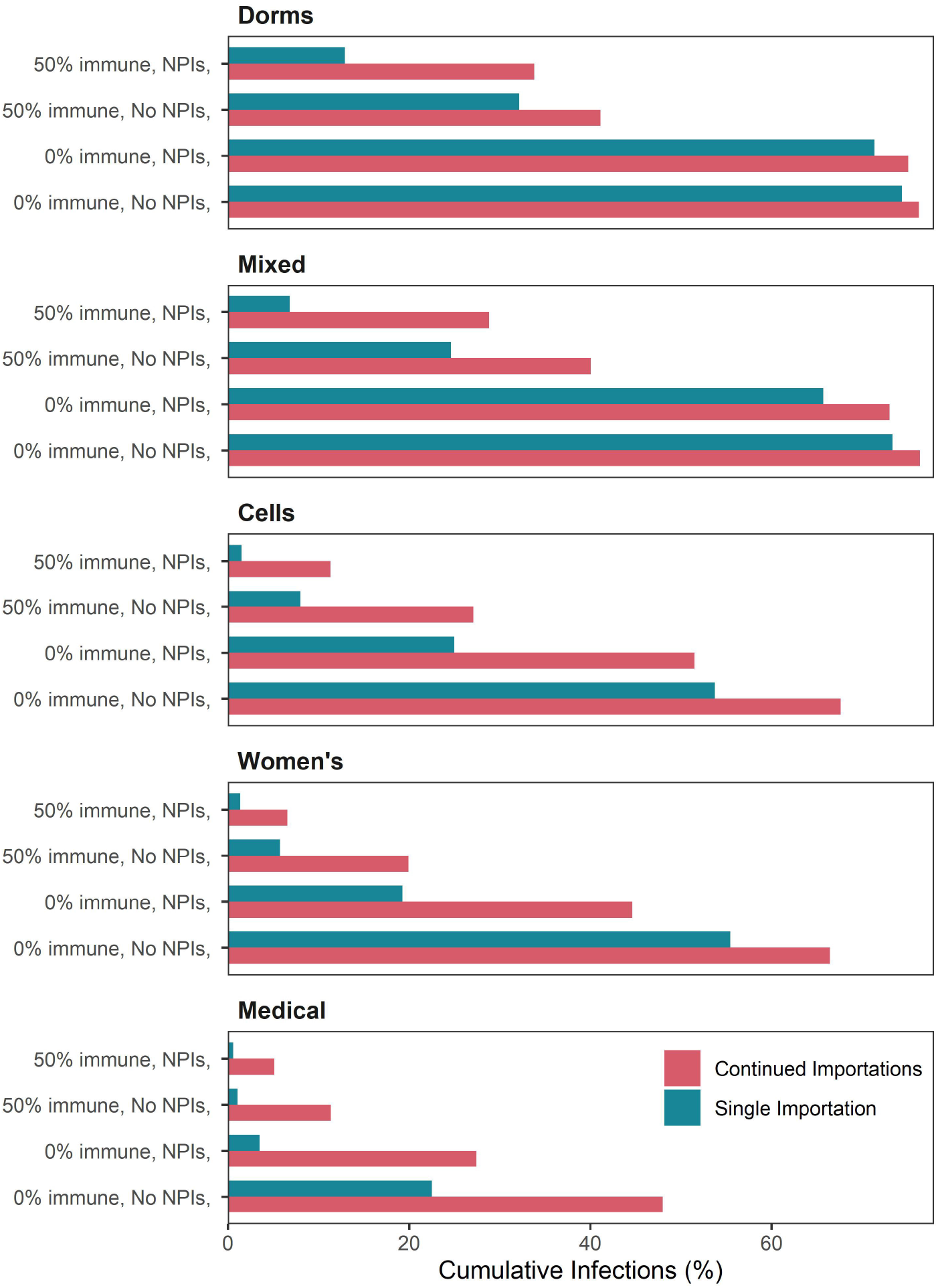
Comparison of cumulative resident infections over 200 days for prisons with resumption of in-person activities and realistic vaccine coverage, varying baseline immunity, NPI usage, and whether variant infection is imported once or repeatedly. Figure shows average cumulative infections among residents across 500 model simulations over 200 days with no importations after day 1 (main analysis; single importation) or 0.1% daily incidence among susceptible staff members (sensitivity analysis; continued importations) for each scenario shown. Results showing the full set of scenarios are available in the appendix (Appendix Figure S19).

Our main analyses assumed that vaccination of both residents and correctional staff occurred prior to the introduction of a new infection. In many states, however, vaccination of incarcerated populations is at an early stage as of the writing of this paper, with a minority of residents vaccinated. Vaccination of residents has substantially less impact when scaled up concurrently with infection introduction, and outbreaks in these settings are expected to be far worse. In settings that are currently at lower risk, such as prisons with cells, moderate to high baseline immunity, and 90% vaccine coverage, outbreaks are twice as large with concurrent vaccination scale-up compared to completed vaccination scale-up at baseline (Appendix Figures S21-S22).

Given limited available data on correctional staff and flatlining of staff coverage in California prisons, we did not focus on analyzing the effects of increasing staff coverage. However, even if staff coverage doubled to 80%, once an infection is already introduced to a prison, this higher coverage was a relatively minor determinant of resident infections and hospitalizations (Appendix Figures S23-S24).

## Discussion

As highly transmissible Covid-19 viral variants proliferate throughout the US, their introduction into the nation’s 110 federal prisons and 1,833 state prisons is inevitable (32). Resumption of in-person activities, while undoubtedly important for the health and welfare of incarcerated people, can also greatly increase the risk and size of outbreaks. Achieving high levels of vaccination prior to resumption of in-person activities lowers risks, as does continuing widespread use of effective NPIs. These findings hold even in prisons that predominantly house residents in lower-occupancy rooms, or that have accrued substantial levels of natural immunity from prior outbreaks.

Resuming in-person activities safely requires a multi-faceted approach. In addition to vaccination and NPIs (e.g., masking) to reduce transmission, other measures may prevent the introduction of infections, limit the extent of spread, or mitigate harms. Sensitivity analyses showed a substantial increase in expected infections and hospitalizations when the likelihood of repeated importation of infections is high (Figure 4), emphasizing the importance of preventing infections from being introduced. While of limited utility for controlling outbreaks once an infection is introduced (Appendix Figure S23), staff vaccination is crucial for choking one of the main avenues of introduction (6,33). Maintaining ongoing screening and testing of residents, staff, and visitors may also prevent introductions and limit the spread of prison outbreaks. Furthermore, analysis of hospitalizations (Figure 3) reveals that older and medically vulnerable residents should receive additional protections beyond vaccine priority, such as lower occupancy housing and additional NPIs. Our study demonstrates the benefits of lower occupancy housing and points to the potential impact of depopulation (16), which other analyses have concluded is an important strategy to reduce transmission (34,35).

The frequency of importation of infections is likely setting-specific given differences in ongoing community epidemics and vaccination coverage. Whether outbreaks more closely resemble our main analysis with a single imported infection (Figure 2) or our sensitivity analysis with repeated importation (Figure 4) depends on setting, future trends in infections, and measures to reduce introductions. Where Covid-19 infection rates in the general population are high or in carceral settings that have large inflows, such as reception centers and jails, repeated importation of infections is more likely, and our main projections (Figure 2) would be underestimates.

Our main results reveal that prioritizing immediate efforts to achieve widespread vaccination among incarcerated people is critical (Figure 2). Our sensitivity analysis shows that if vaccination scale-up is delayed and an infection is introduced in the meantime, the rate of spread is likely to outpace feasible vaccine scale-up, especially in prisons that have resumed in-person activities (Appendix Figure S21). Currently, vaccine coverage and acceptance among incarcerated people and correctional staff is highly variable across prison systems (17–19). Employing the best-available vaccination offer and re-offer strategies may be required to increase acceptance especially among those who perceive their risk to be low or are distrustful of prison authorities or healthcare providers (19,36).

Regular surveillance testing arms prison health officials with early knowledge of the presence and spread of infections but, to our knowledge, most correctional systems are not using genetic sequencing technology to surveil for the presence of variants. Hence, detection of an outbreak cannot indicate whether the additional precautions needed to curb the spread of a variant of concern are necessary. Alternatively, the seeding of wild type virus may not warrant complete shutdown (Appendix Figure S17-S18). Until prisons can rapidly distinguish between wild type infections and variants of concern, detecting new infections may trigger policies that under- or over-protect residents and staff.

Our study has several limitations, First, our results may not be fully generalizable to other carceral settings. The impacts of infection introduction on incarcerated populations depend in part on how likely prisons are to detect outbreaks and take measures to isolate and quarantine. Our results are based on modeling of widespread testing as practiced in California’s prisons during 2020-21. However, in settings where screening and testing are less intensive, the risk and size of outbreaks are likely to be larger than our estimates. Although we modeled a range of prison types, incarcerated populations, and levels of prior immunity, outcomes in individual prisons could differ if their housing configurations or demographics diverge substantially from those modeled. Second, while we considered variant scenarios that broadly represent variants like *B.1.1.7* along with scenarios considering introduction of wild type variants, other variants are currently circulating globally, and new variants are emerging even as the scientific community works to keep pace in characterizing them epidemiologically (12,15). Some variants may be able to completely evade natural immunity from prior wild type infection; although we did not model these explicitly, we did model equivalent scenarios with 0% baseline immunity. However, variants that are simultaneously able to avoid vaccine-induced immunity and much more likely to cause severe outcomes, though not yet detected, would require further evaluation.

Ultimately, prison health authorities will need to determine to what extent the risks of infection and serious Covid-19 outcomes must be reduced before the benefits of resumption of in-person activities outweigh the risks. While vaccination reduces risks of serious Covid-19 outcomes, the longer-term implications of even mild infections themselves are not yet well understood. Published studies have detailed long-term sequelae from both mild and severe Covid-19 infections, including respiratory conditions, cardiovascular disease, and mental health disorders (37,38). Furthermore, outbreaks in prisons can have consequences for the broader community and uncontrolled spread of the virus can lead to further mutations that produce even harder to combat variants, as has been observed for other infectious diseases (39,40). Decision-making regarding pandemic control should be integrated between public health authorities in both incarcerated and free-living settings. Neglecting prison health and deprioritizing vaccination is both unethical and bad policy.

Our analysis yields important policy conclusions based on models that incorporate detailed primary data from the second-largest state prison system in the United States, explicitly reflect demography, residential structure, and mixing over a range of prison types, and can reproduce the size and heterogeneity of viral spread reported in the literature. In summary, to enable resumption of in-person activities so important for the health and wellbeing of incarcerated people, prisons should prioritize widespread vaccination of their populations as soon as possible, remain dedicated to NPIs and regular surveillance testing including genetic sequencing, and continue to protect vulnerable populations.

## Data Availability

The mathematical model used in the manuscript is parameterized by data from public sources and non-publicly available data obtained via a Data Use Agreement between Stanford University and the California Department of Corrections and Rehabilitation.

## Acknowledgements

We thank staff members at the California Department of Corrections and Rehabilitation for providing data and assistance with interpretation of study results. We would also like to thank Liesl Hagan, MPH at the United States Centers for Disease Control and Prevention and Robert Canning, PhD at the California Department of Corrections and Rehabilitation for sharing their expertise to help inform our selection of prisons for this analysis. We also acknowledge assistance from other members of the SC-COSMO consortium.

## References

1. Saloner B, Parish K, Ward JA, DiLaura G, Dolovich S. COVID-19 Cases and Deaths in Federal and State Prisons. JAMA. 2020 Aug 11;324(6):602.

2. Toblin RL, Hagan LM. COVID-19 Case and Mortality Rates in the Federal Bureau of Prisons. American Journal of Preventive Medicine [Internet]. 2021 Feb 24 [cited 2021 Mar 10];0(0). Available from: https://www.ajpmonline.org/article/S0749-3797(21)00119-7/abstract

3. Burkhalter E, Colón I, Derr B, Gamio L, Griesbach R, Klein AH, et al. Incarcerated and Infected: How the Virus Tore Through the U.S. Prison System. The New York Times [Internet]. 2021 Apr 10 [cited 2021 Apr 14]; Available from: https://www.nytimes.com/interactive/2021/04/10/us/covid-prison-outbreak.html

4. Committee on the Best Practices for Implementing Decarceration as a Strategy to Mitigate the Spread of COVID-19 in Correctional Facilities. Decarcerating Correctional Facilities during COVID-19: Advancing Health, Equity, and Safety [Internet]. Wang EA, Western B, Backes EP, Schuck J, editors. The National Academies Press; [cited 2021 Mar 10]. Available from: https://www.nap.edu/read/25945/chapter/1

5. Chin ET, Ryckman T, Prince L, Leidner D, Alarid-Escudero F, Andrews JR, et al. Covid-19 in the California State Prison System: An Observational Study of Decarceration, Ongoing Risks, and Risk Factors. medRxiv. 2021 Mar 8;2021.03.04.21252942.

6. Hagan LM. Mass Testing for SARS-CoV-2 in 16 Prisons and Jails — Six Jurisdictions, United States, April–May 2020. MMWR Morb Mortal Wkly Rep [Internet]. 2020 [cited 2021 Apr 6];69. Available from: https://www.cdc.gov/mmwr/volumes/69/wr/mm6933a3.htm

7. After rampant COVID cases and mass vaccines, is California’s prison system nearing ‘herd immunity’? [Internet]. The Mercury News. 2021 [cited 2021 Apr 27]. Available from: https://www.mercurynews.com/2021/03/07/after-rampant-covid-cases-and-mass-vaccines-is-california-prison-system-nearing-herd-immunity

8. California prisons had so many COVID cases, they now have herd immunity. More could follow. The Sacramento Bee [Internet]. [cited 2021 Apr 27]; Available from: https://www.sacbee.com/news/politics-government/the-state-worker/article249734353.html

9. Aschwanden C. The false promise of herd immunity for COVID-19. Nature. 2020 Oct 21;587(7832):26–8.

10. Keeling MJ, Rohani P. Modeling Infectious Diseases in Humans and Animals [Internet]. Princeton University Press; 2007 [cited 2021 Mar 19]. Available from: https://press.princeton.edu/books/hardcover/9780691116174/modeling-infectious-diseases-in-humans-and-animals

11. Moore JP, Offit PA. SARS-CoV-2 Vaccines and the Growing Threat of Viral Variants. JAMA. 2021 Mar 2;325(9):821–2.

12. CDC. SARS-CoV-2 Variant Classifications and Definitions [Internet]. Centers for Disease Control and Prevention. 2021 [cited 2021 Mar 23]. Available from: https://www.cdc.gov/coronavirus/2019-ncov/cases-updates/variant-surveillance/variant-info.html

13. Graham MS, Sudre CH, May A, Antonelli M, Murray B, Varsavsky T, et al. Changes in symptomatology, reinfection, and transmissibility associated with the SARS-CoV-2 variant B.1.1.7: an ecological study. The Lancet Public Health. 2021 May 1;6(5):e335–45.

14. Leatherby L, Reinhard S. More Contagious Variant Is Spreading Fast in U.S., Even as Overall Cases Level Off. The New York Times [Internet]. 2021 Mar 6 [cited 2021 Mar 10]; Available from: https://www.nytimes.com/interactive/2021/03/06/us/coronavirus-variant-sequencing.html

15. Mullen JL, Tsueng G, Latif AA, Alkuzweny M, Cano M, Haag E, et al. Outbreak.info: a standardized, open-source database of COVID-19 resources and epidemiology data [Internet]. outbreak.info. [cited 2021 Apr 28]. Available from: https://outbreak.info/

16. Barsky BA, Reinhart E, Farmer P, Keshavjee S. Vaccination plus Decarceration — Stopping Covid-19 in Jails and Prisons. New England Journal of Medicine. 2021 Apr 29;384(17):1583–5.

17. Quandt KR, Prison Policy Initiative. Incarcerated people and corrections staff should be prioritized in COVID-19 vaccination plans [Internet]. 2021 [cited 2021 Mar 25]. Available from: https://www.prisonpolicy.org/blog/2020/12/08/covid-vaccination-plans/

18. UCLA Law COVID-19 Behind Bars Data Project [Internet]. [cited 2021 Apr 28]. Available from: https://uclacovidbehindbars.org/

19. Chin ET, Leidner D, Ryckman T, Liu Y, Prince L, Alarid-Escudero F, et al. Covid-19 Vaccine Acceptance among Residents of California State Prisons. Forthcoming. 2021 Mar;

20. He X, Lau EHY, Wu P, Deng X, Wang J, Hao X, et al. Temporal dynamics in viral shedding and transmissibility of COVID-19. Nature Medicine. 2020 May;26(5):672–5.

21. Lauer SA, Grantz KH, Bi Q, Jones FK, Zheng Q, Meredith HR, et al. The Incubation Period of Coronavirus Disease 2019 (COVID-19) From Publicly Reported Confirmed Cases: Estimation and Application. Ann Intern Med [Internet]. 2020 Mar 10 [cited 2021 Mar 18]; Available from: https://www.ncbi.nlm.nih.gov/pmc/articles/PMC7081172/

22. Ashcroft P, Huisman JS, Lehtinen S, Bouman JA, Althaus CL, Regoes RR, et al. COVID-19 infectivity profile correction. Swiss Medical Weekly [Internet]. 2020 Aug 5 [cited 2021 Mar 18];150(3132). Available from: https://smw.ch/article/doi/smw.2020.20336

23. Verity R, Okell LC, Dorigatti I, Winskill P, Whittaker C, Imai N, et al. Estimates of the severity of coronavirus disease 2019: a model-based analysis. The Lancet Infectious Diseases. 2020 Jun 1;20(6):669–77.

24. Tuite AR, Fisman DN, Odutayo A, Bobos P, Allen V, Bogoch II, et al. COVID-19 Hospitalizations, ICU Admissions and Deaths Associated with the New Variants of Concern [Internet]. Ontario COVID-19 Science Advisory Table. [cited 2021 Apr 8]. Available from: https://covid19-sciencetable.ca/sciencebrief/covid-19-hospitalizations-icu-admissions-and-deaths-associated-with-the-new-variants-of-concern/

25. Amit S, Regev-Yochay G, Afek A, Kreiss Y, Leshem E. Early rate reductions of SARS-CoV-2 infection and COVID-19 in BNT162b2 vaccine recipients. The Lancet. 2021 Mar 6;397(10277):875– 7.

26. Polack FP, Thomas SJ, Kitchin N, Absalon J, Gurtman A, Lockhart S, et al. Safety and Efficacy of the BNT162b2 mRNA Covid-19 Vaccine. New England Journal of Medicine. 2020 Dec 31;383(27):2603–15.

27. Kucirka LM, Lauer SA, Laeyendecker O, Boon D, Lessler J. Variation in False-Negative Rate of Reverse Transcriptase Polymerase Chain Reaction-Based SARS-CoV-2 Tests by Time Since Exposure. Ann Intern Med. 2020 Aug 18;173(4):262–7.

28. Staphan J. Census of State and Federal Correctional Facilities, 2005 [Internet]. Bureau of Justice Statistics; Available from: https://www.bjs.gov/content/pub/pdf/csfcf05.pdf

29. US Census Bureau. American Community Survey Data [Internet]. The United States Census Bureau. [cited 2021 Mar 9]. Available from: https://www.census.gov/programs-surveys/acs/data.html

30. US Centers for Disease Control and Prevention. Life Tables [Internet]. 2019 [cited 2021 Mar 9]. Available from: https://www.cdc.gov/nchs/products/life_tables.htm

31. Stout NK, Goldie SJ. Keeping the Noise Down: Common Random Numbers for Disease Simulation Modeling. Health Care Manag Sci. 2008 Dec;11(4):399–406.

32. Sawyer W, Wagner P. Mass Incarceration: The Whole Pie 2020 [Internet]. 2020 [cited 2021 Apr 15]. Available from: https://www.prisonpolicy.org/reports/pie2020.html

33. Wallace M, James AE, Silver R, Koh M, Tobolowsky FA, Simonson S, et al. Rapid Transmission of Severe Acute Respiratory Syndrome Coronavirus 2 in Detention Facility, Louisiana, USA, May– June, 2020 - Volume 27, Number 2—February 2021 - Emerging Infectious Diseases journal - CDC. Emerging Infectious Diseases [Internet]. 2021 Feb [cited 2021 Apr 26];27(2). Available from: https://www.nc.cdc.gov/eid/article/27/2/20-4158_article

34. Malloy GSP, Puglisi L, Brandeau ML, Harvey TD, Wang EA. Effectiveness of interventions to reduce COVID-19 transmission in a large urban jail: a model-based analysis. BMJ Open. 2021 Feb 1;11(2):e042898.

35. Vest N, Johnson O, Nowotny K, Brinkley-Rubinstein L. Prison Population Reductions and COVID-19: A Latent Profile Analysis Synthesizing Recent Evidence From the Texas State Prison System. J Urban Health. 2021 Feb 1;98(1):53–8.

36. Stern MF. Willingness to Receive a COVID-19 Vaccination Among Incarcerated or Detained Persons in Correctional and Detention Facilities — Four States, September–December 2020. MMWR Morb Mortal Wkly Rep [Internet]. 2021 [cited 2021 Apr 28];70. Available from: https://www.cdc.gov/mmwr/volumes/70/wr/mm7013a3.htm

37. Al-Aly Z, Xie Y, Bowe B. High-dimensional characterization of post-acute sequalae of COVID-19. Nature. 2021 Apr 22;1–8.

38. CDC. Post-COVID Conditions [Internet]. Centers for Disease Control and Prevention. 2020 [cited 2021 Apr 26]. Available from: https://www.cdc.gov/coronavirus/2019-ncov/long-term-effects.html

39. Mabud TS, de Lourdes Delgado Alves M, Ko AI, Basu S, Walter KS, Cohen T, et al. Evaluating strategies for control of tuberculosis in prisons and prevention of spillover into communities: An observational and modeling study from Brazil. PLoS Med. 2019 Jan;16(1):e1002737.

40. Sacchi FPC, Praça RM, Tatara MB, Simonsen V, Ferrazoli L, Croda MG, et al. Prisons as reservoir for community transmission of tuberculosis, Brazil. Emerg Infect Dis. 2015 Mar;21(3):452–5.

