## Appendix for "Outbreaks of Covid-19 Variants in Prisons: A Mathematical Modeling Analysis of Vaccination and Re-Opening Policies"

**Supplemental Appendix for Outbreaks of Covid-19 Variants in Prisons: A Mathematical Modeling Analysis of Vaccination and Re-Opening Policies**

Theresa Ryckman, Elizabeth T. Chin, Lea Prince, David Leidner, Elizabeth Long, David M. Studdert, Joshua A. Salomon, Fernando Alarid-Escudero, Jason R. Andrews, Jeremy D. Goldhaber-Fiebert

**Microsimulation Model**

The transmission-dynamic stochastic microsimulation model follows individual residents and correctional staff within a prison. The model reflects the prison’s residential structure and we simulate different prisons by instantiating the model with prison-specific characteristics.

Each day, for each incarcerated person and correctional staff member who is currently susceptible to infection, we compute the risk of infection using a set of transmission equations.

*Transmission Equations*

Equation 1 shows the transmission rate for resident $i$who lives in room $r$, building$b$, and yard $y$and who interacts with:

- Other residents, $res$, in the same room $r$(e.g., cell or dorm)
- Other residents in the same building $b$
- If individual $i$participates in activities $a,$(labor $l$, school $s$, and/or other activities $o$), then he or she also interacts with other residents in the same yard $y$who participate in labor, school, and/or other activities, with participation in each activity type noted with an indicator function $1\left\{ . \right\}$ in Equation 1
- Correctional staff, $cs$, in the same prison

$$\left[ 1 \right] {rate}_{i}={rate}_{r}N_{eff,r}\left( \frac{I_{r}}{N_{r}} \right)+rate_{b}N_{eff,b}\left( \frac{I_{b}}{N_{b}} \right)+1\left\{ labor \right\}{rate}_{a}N_{l,y}\left( \frac{I_{l,y}}{N_{l,y}} \right)+1\left\{ school \right\}{rate}_{a}N_{s,y}\left( \frac{I_{s,y}}{N_{s,y}} \right)+1\left\{ other activities \right\}{rate}_{a}N_{o,y}\left( \frac{I_{o,y}}{N_{o,y}} \right)+\beta_{res,cs}\left( \frac{I_{cs}}{N_{cs}} \right)$$

Equation 2 shows the transmission rate for correctional staff member *j,* who interacts with:

- Residents, $res$, in the same prison
- Other correctional staff, $cs$, in the same prison

$$\left[ 2 \right] rate_{j}=\beta_{cs,res}\left( \frac{I_{res}}{N_{res}} \right)+\beta_{cs,cs}\left( \frac{I_{cs}}{N_{cs}} \right)$$

In both equations:

- $rate$denotes the rate of infection per infected contact for each type of contact (room, building, or activities)
- $\beta$ denotes the effective contact rate (between residents and correctional staff, correctional staff and residents, and correctional staff and other correctional staff)
- $N$ denotes the total number of individuals
- $I$denotes the total number of infectious individuals
- $N_{eff}$denotes the number of effective contacts, in cases where it differs from the total number of individuals (room and building contacts only, details in next section)

*Effective Contacts and Model Calibration*

Calibrated parameters are summarized in Table S1 and described in more detail below.

**Table S1: Calibrated transmission parameters**

| Parameter | Abbreviation | Value |
| --- | --- | --- |
| Daily rate of infection per infectious room contact | ${rate}_{r}$ | 0.05 |
| Daily rate of infection per infectious building contact | ${rate}_{b}$ | 0.005 |
| Number of effective room contacts | $N_{eff,r}$ | $\min\left\{ \sum_{i=0}^{N_{r}-3} \frac{1}{\left( 1.05 \right)^{i}}, N_{r} \right\}-1$ |
| Number of effective building contacts | $N_{eff,b}$ | All rooms except medium dorms: $N_{b}$  Medium dorms: $\min\{100, N_{b}\}$ |
| Daily rate of infection per infectious activity contact | $rate_{a}$ | 0.025 |
| Effective contact rate between residents and correctional staff | $\beta_{res,cs}$, $\beta_{cs,res}$ | 0.05 |
| Effective contact rate between correctional staff | $\beta_{cs,cs}$ | 0.15 |

The rate of infection per infectious room contact, ${rate}_{r}$, the rate of infection per infectious building contact, ${rate}_{b},$and the relationships between the room or building censuses, $N_{r}$ and $N_{b}$, and the effective room or building contacts, $N_{eff,r}$and $N_{eff,b}$ , were calibrated to empirical estimates of the prison within-room secondary attack rate (SAR) among residents across California state prisons, which we estimated from primary CDCR data. We categorized rooms with at least two residents into four types: double cells (2 occupants), small dorms (3-10 occupants), medium dorms (11-30 occupants), and large dorms (31 or more occupants). We simulated outbreaks in prisons with different room types and calculated the observed SAR from the model-predicted output (i.e., the number of subsequent detected secondary cases among roommates of the first case detected in a room, within 14 days of the detection of the index case, divided by the number of occupants in that room), stratified by room type. We repeated these simulations across multiple combinations of values of ${rate}_{r}$ and ${rate}_{b}$ and functional forms for $N_{eff,r}$ and $N_{eff,b}$.

In many contexts, the number of contacts people have does not grow with constant proportionality to the total population size which is why many epidemic models use transmission terms like $\beta\frac{SI}{N}$ instead of $\beta SI$ (1–3). In dormitories with tens or even hundreds of occupants, any individual may have contact with a fraction of the total occupants in any given day (in contrast to 2 person cells). Hence, we selected various functional forms for $N_{eff,r}$ to allow for effective contacts to rise more slowly than room census. Further informing this choice was that the empirical estimates of SAR do not strictly increase with room census.

The number of effective building contacts, $N_{eff,b}$, was generally selected to be equal to building census (minus room census), except that we capped $N_{eff,b}$ for medium dorms (11-30 occupants). While there are generally few large dorms per building in CDCR prisons, there are often many medium dorms in one building, yielding a large number of building contacts if uncapped. However, those living in medium (and large) dorms are likely to interact more with their roommates and less with others in the building since bathrooms, recreational rooms, and other common facilities are often specific to a dorm and not shared across dorms.

We compared the model-predicted SAR to corresponding empirical estimates of SAR, and selected parameters which yielded model output that fell within the 95% confidence bounds of the empirical estimates across room types (Figure S1). Based on these simulations, we selected a daily transmission rate per infected room contact of 0.05 (which roughly corresponds to a transmission probability of 5%) and a daily transmission rate per infected building contact of 0.005 (which roughly corresponds to a transmission probability of 0.05%). We capped effective building contacts for medium dorms at 100 contacts and established the following functional form for effective room contacts (Table S1, equation 3, Figure S2):

[3]	$N_{eff,r_{i}}=\min\left\{ 4+\sum_{i=0}^{{(N}_{r_{i}}-1)-4} \frac{1}{\left( 1.1 \right)^{i}}, {(N}_{r_{i}}-1) \right\}$

**Figure S1: Fit of modeled secondary attack rate to empirical estimates, with contact dampening**

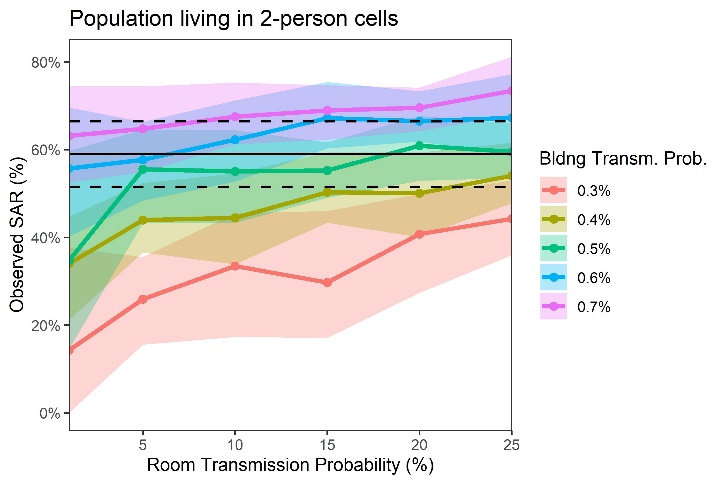

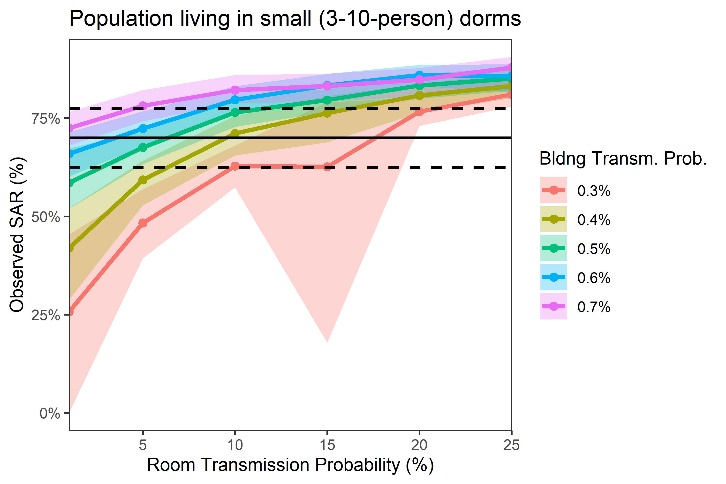

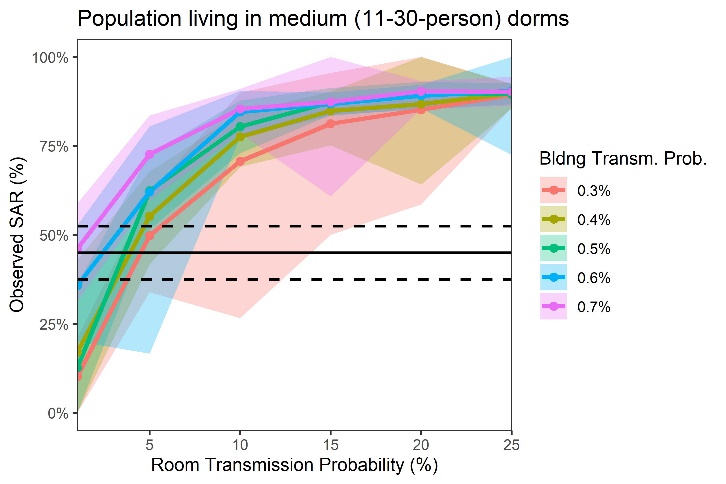

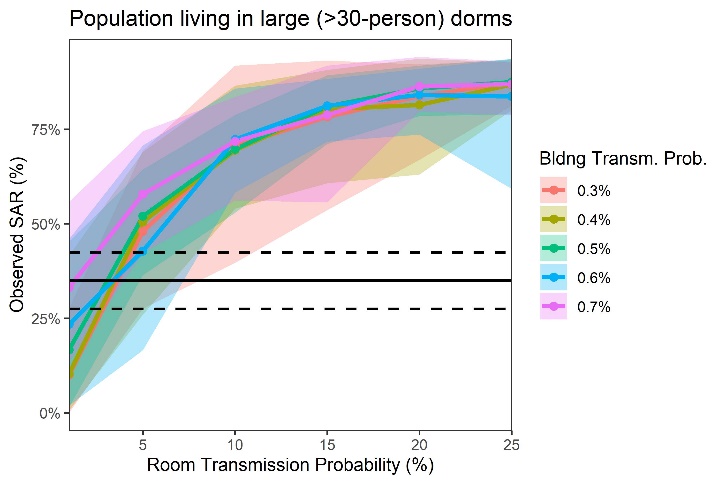

Empirical estimates are shown in black (solid lines), with confidence intervals shown via dashed lines. The 0.5^th^ and 99.5^th^ percentiles on modeled output are shown via colored shading, while the means are shown via colored lines.

**Figure S2: Calibrated relationship between room census and effective room contacts**

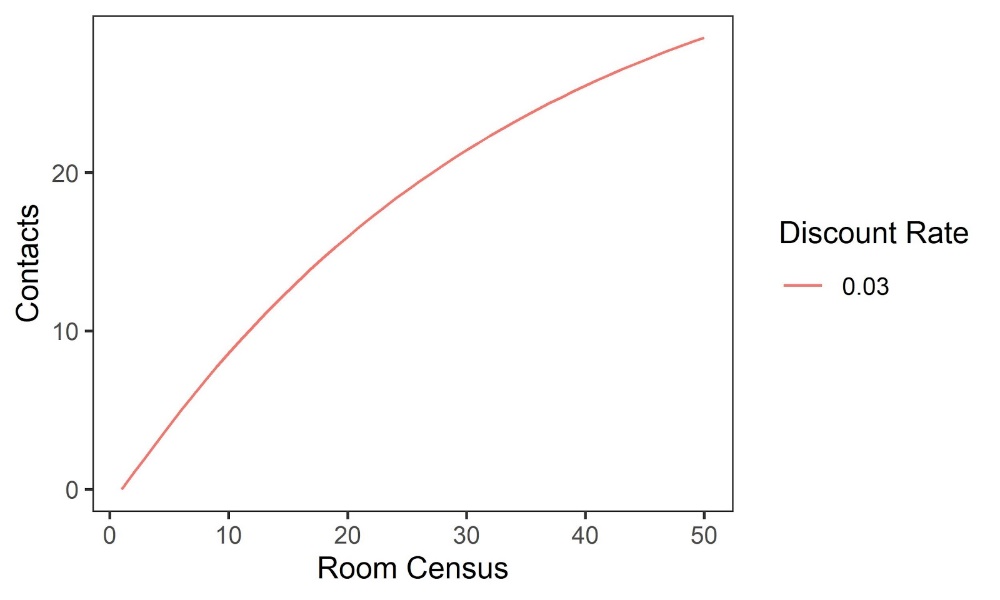

The remaining transmission-related parameters ($rate_{a}, \beta_{res,cs}, \beta_{cs,res},$and $\beta_{cs,cs}$) were selected after these initial four parameters (${rate}_{r}, {rate}_{b}, N_{eff,r},$ and $N_{eff,b})$were calibrated, since they were found to have relatively little influence on the within-room SAR. Transmission from activity-specific contacts and between residents and correctional staff are important in the model because they cause outbreaks that start in one building to spread to additional buildings and yards. This follows from considering a scenario in which an outbreak has already taken off in one part of the prison. In such a case, it can spread elsewhere in the prison even if the activity transmission rate and effective contact rates are minimal because it only requires one activity contact or staff member to be infected/infectious and then transmit to someone else from another part of the prison for the outbreak to then spread through building and room contacts in that other area. We selected the effective contact rates between staff and between staff and residents so that the average infected staff person would infect one other staff member and 1-2 residents (Figure S3; $\beta_{res,cs},=\beta_{cs,res}=0.05$and $\beta_{cs,cs}=0.15$)), implying a basic reproductive number on the high end of estimates from free living populations (4,5). We selected the rate of transmission per infected labor/school/other activity contact, $rate_{a}=0.025$, which is half of the room transmission rate, to reflect the fact that activity contacts are likely of shorter duration and proximity than in-room contacts, especially with non-pharmaceutical interventions such as masking and attempts to physically distance are in place at activities.

**Figure S3: Calibrated secondary infections from one index staff infection**

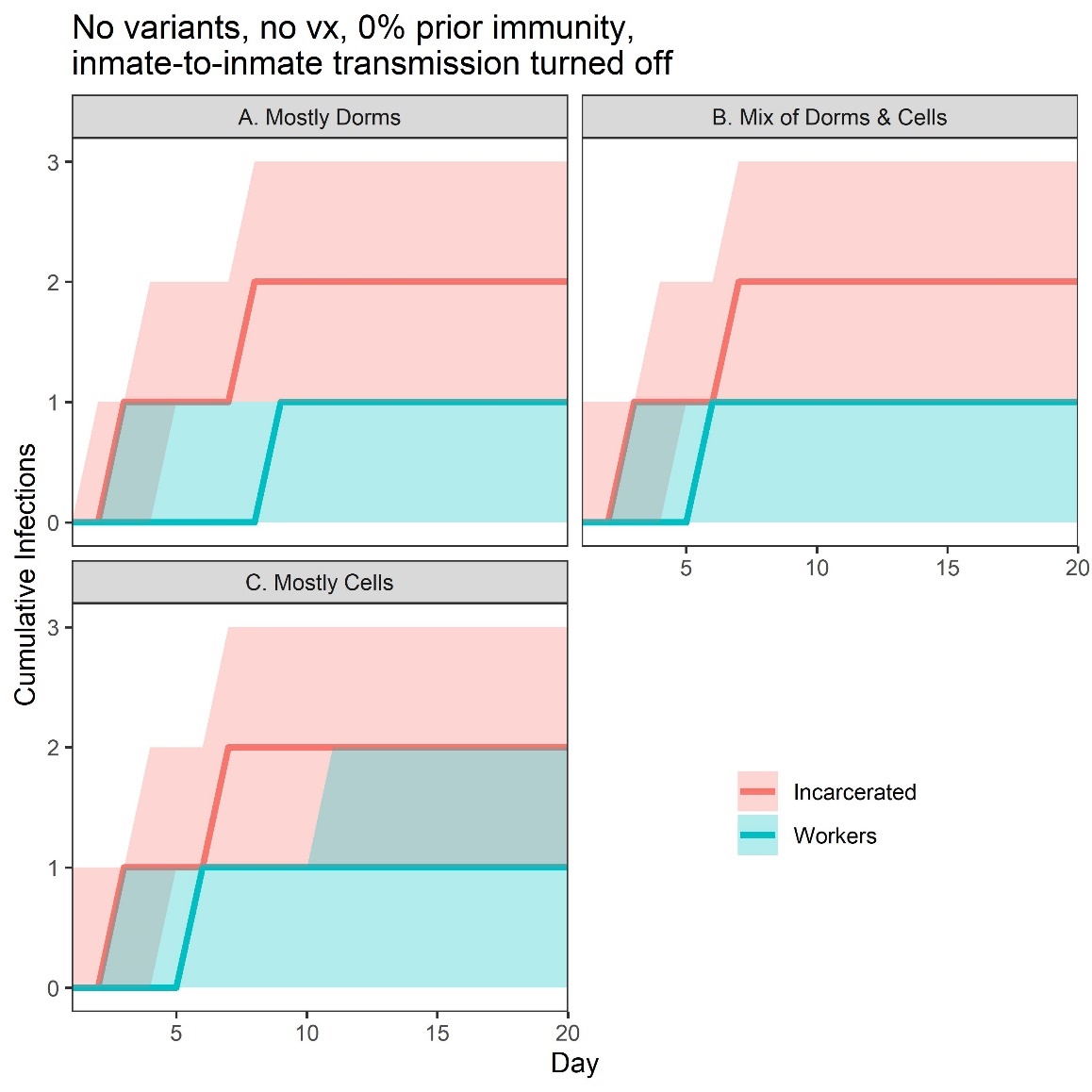

The figure shows the results of seeding one staff member infected with wild-type Covid-19 in a prison with no vaccination and no baseline immunity to infection. We simulated for 20 days, with all transmission between residents disabled to focus on staff-related transmission. Medians with interquartile ranges are shown.

*Modeling the residential arrangement of prisons*

Our model replicates a prison’s *residential arrangement*, including the number of rooms by occupancy, within buildings, and in turn within yards. This allows the analysis to simulate the spread of infections across a prison (Figures S4-S5).

**Figure S4: Simulated spread of Covid-19 infections in a prison with mostly dorms**

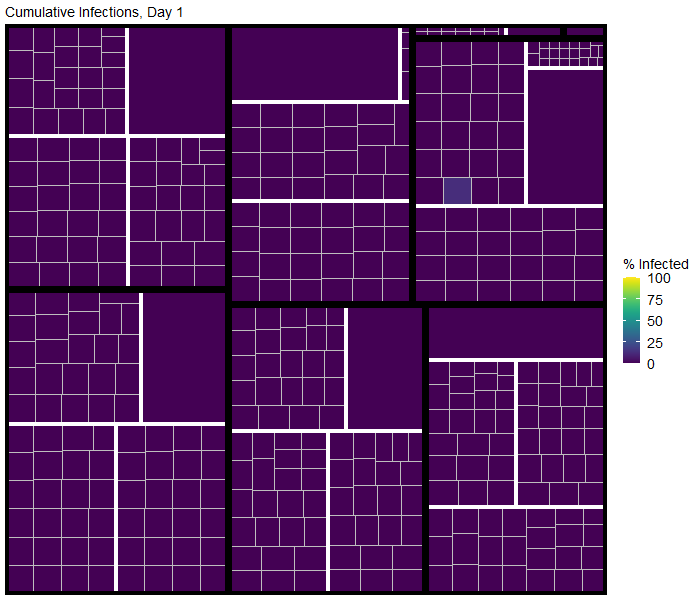

In this figure, thick black boxes designate yards, thick white boxes designate buildings within yards, and thin white boxes designate rooms within buildings. The number of occupants scales the sizes of the rooms. Rooms are shaded by the percentage of those living in the room who have been infected with Covid-19 over time.

**Figure S5: Simulated spread of Covid-19 infections in a prison with mostly cells**

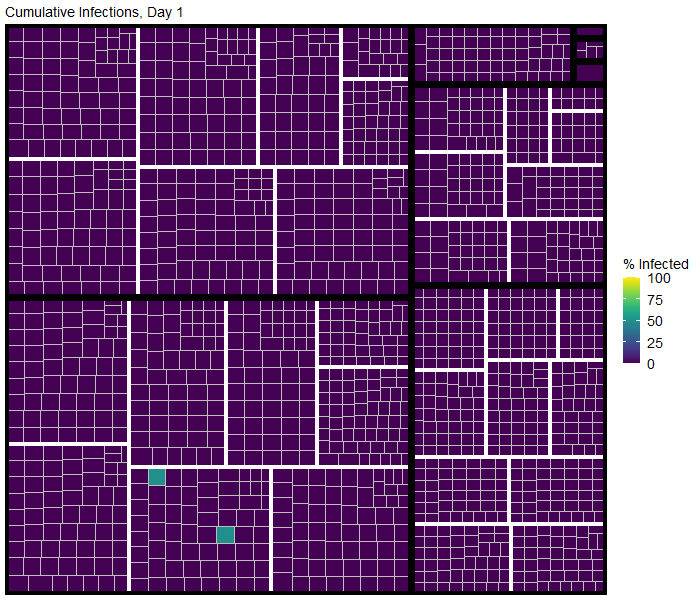

In this figure, thick black boxes designate yards, thick white boxes designate buildings within yards, and thin white boxes designate rooms within buildings. The number of occupants scales the sizes of the rooms. Rooms are shaded by the percentage of those living in the room who have been infected with Covid-19 over time.

*Testing, Quarantine, and Isolation*

Based on CDCR policy, we model four main types of case detection among residents:

1. Background surveillance testing: CDCR developed a Covid-19 risk score to grade each resident’s probability of severe health outcomes following Covid-19 infection. Scores correspond to the presence of demographic and clinical characteristics identified in the scientific literature as risk factors for severe Covid-19-related illness (e.g., age >65 years, immunocompromised) (Table S2) (6). Residents are flagged as high-risk if their Covid-19 risk score is at least 4. Some prisons house older and more medically vulnerable individuals than others. If a prison houses fewer than 60 high-risk residents, we model testing for all high-risk residents every 2 weeks. Otherwise, a random selection of 15 high-risk residents in each yard is tested every 2 weeks.
2. Reactive testing: When a case is detected (e.g., via surveillance testing), contacts of the infected resident are also tested. These contacts include everyone in the same room as the detected individual as well as 20% of residents in the same building (but different rooms). If reactive testing identifies additional cases, this could trigger outbreak-level testing (see below).
3. Outbreak-level testing: A single case will only trigger reactive testing. However, if 2 or more cases are detected in the prison within a 14-day window, outbreak-level testing is triggered for all yards in which cases were detected. If all detected cases were in the same building, 80% of the residents of that building are tested. If cases were detected in multiple buildings in the same yard, 80% of the residents of that yard are tested.
4. Testing of hospitalized patients: All infections that are severe enough to require hospitalization are detected and subsequently tested. Regardless of test result (i.e. even if the result is a false negative), these severely infected individuals are isolated and hospitalized.

All detected cases are isolated for 14 days. All residents of a building with a detected case are put into quarantine, during which they continue to interact with others in the same building and with staff but they no longer interact with any residents outside of their building (e.g., via activities). Quarantine lasts for 14 days after the last day on which a case was detected. For buildings undergoing large outbreaks, quarantine could therefore last far longer than 14 days. We include a 4-day lag between the day a test sample is taken and the day a detected case is isolated, and their building quarantined.

Detection of staff infections is modeled in a simplified way. Correctional staff infections can only be detected via symptom screening. Every staff member is screened daily. Symptomatic infected staff (regardless of infectious status) have a 50% chance of being detected for each day they are symptomatic. Asymptomatic infected staff have a 0% chance of being detected. Detected staff isolate for 14 days from the day of detection.

**Table S2: Covid-19 risk score criteria**

| **Condition** | **Definition** | **Weighted Score** |
| --- | --- | --- |
| Age 65+ | Chronologic age of 65 years or above | 4 |
| Advanced liver disease | Has advanced liver disease (cirrhosis/end stage liver disease) | 2 |
| Asthma | Persistent asthma (moderate or severe) as defined by the  California Correctional Health Care Services (CCHCS) asthma condition specifications | 1 |
| Cancer | High risk cancer as defined by the CCHCS cancer condition specifications (excludes most diagnoses of skin cancer and “personal history of” cancers”) | 2 |
| Chronic Lung Disease (other) | Has cystic fibrosis, pneumoconiosis, or pulmonary fibrosis | 1 |
| Chronic Obstructive Pulmonary Disease (COPD) | Has Chronic Obstructive Pulmonary Disease | 2 |
| Cardiovascular Disease (CVD) | Has any of the following:” cerebrovascular disease, congestive heart failure, congenital heart disease, ischemic heart disease, peripheral vascular disease, thromboembolic disease, and valvular disease | 1 |
| Cardiovascular Disease (CVD; high risk) | Is high risk for any of the following:” cerebrovascular disease, congestive heart failure, congenital heart disease, ischemic heart disease, peripheral vascular disease, thromboembolic disease, and valvular disease | 1 |
| Diabetes | Has diabetes | 1 |
| Diabetes (high risk) | Meets the criteria for high risk diabetes as defined by the CCHS diabetes condition specifications | 1 |
| HIV | Has HIV | 1 |
| HIV (poorly controlled) | Has HIV with a CD4 count < 200 | 1 |
| Immunocompromised | Has any of the following: aplastic anemia, histiocytosis, immunosuppressed, organ transplant, other transplant | 2 |
| Morbid Obesity | Body Mass Index of 40 or above | 1 |
| Other Chronic Conditions | Has any of the following with a high-risk rating: hypertension, coccidiomycosis, connective tissue disorder, dementia/Parkinson’s disease, endocrine disorder, multiple sclerosis, myasthenia gravis, neurologic disorder, vasculitis | 1 |
| On Dialysis | On hemodialysis | 2 |
| Pregnant | Actively pregnant | 1 |

Source: Chin et al. (6)

**Prisons**

In the main text, we present analyses for five prisons: a minimum-to-medium security men’s prison in which most residents are housed in dormitories (“dorms”), a maximum-security men’s prison in which most residents are housed in cells (“cells”), a medium-security men’s prison that includes a mix of residents living in a mix of celled housing and dormitories (“mixed”), a medical prison that houses male residents requiring special medical care who tend to be older and sicker than residents in the main prisons (“medical”), and a women’s prison that combines female residents across security levels in yards of varying security levels (“women’s”, this prison has mostly cells and also has a smaller population). Details on the residential arrangement and populations of these prisons are shown in Figure S6-S11.

These figures show percentages of residents by various characteristics. The security level is based on CDCR’s security level system, where 1 is the lowest security level, and 4 is the highest security level. Room sizes are characterized by the number of occupants and include single cells (1 occupant), double cells (2 occupants), small to medium dorms (3-30 occupants), large dorms (31+ occupants). Comorbidities include advanced liver disease, asthma, cancer, COPD, chronic lung disease, cardiovascular disease, diabetes, HIV, immunocompromised, kidney disease (e.g., on dialysis). The activities panel shows the percent of residents that participated in any of the three types of activities out of their rooms (but still in the prison) in the past week with at least 1 other resident. On this panel, “Before Closures” indicates January 2020 (before closures due to Covid-19), and “With Closures” indicates November 2020 (with closures to due Covid-19 implemented). “Labor” includes both jobs that support the upkeep of the prison (resident workers at medical prisons, laundry, kitchen duty, etc.) and industries. “School” includes any educational activities. These are all set to 0% in the “post” period because CDCR has had residents participate in educational activities in their rooms to minimize transmission. “Other” includes several additional activities, including medical appointments, group therapy, and worship.

**Figure S6: Low-to-medium security prison with mostly dorms**

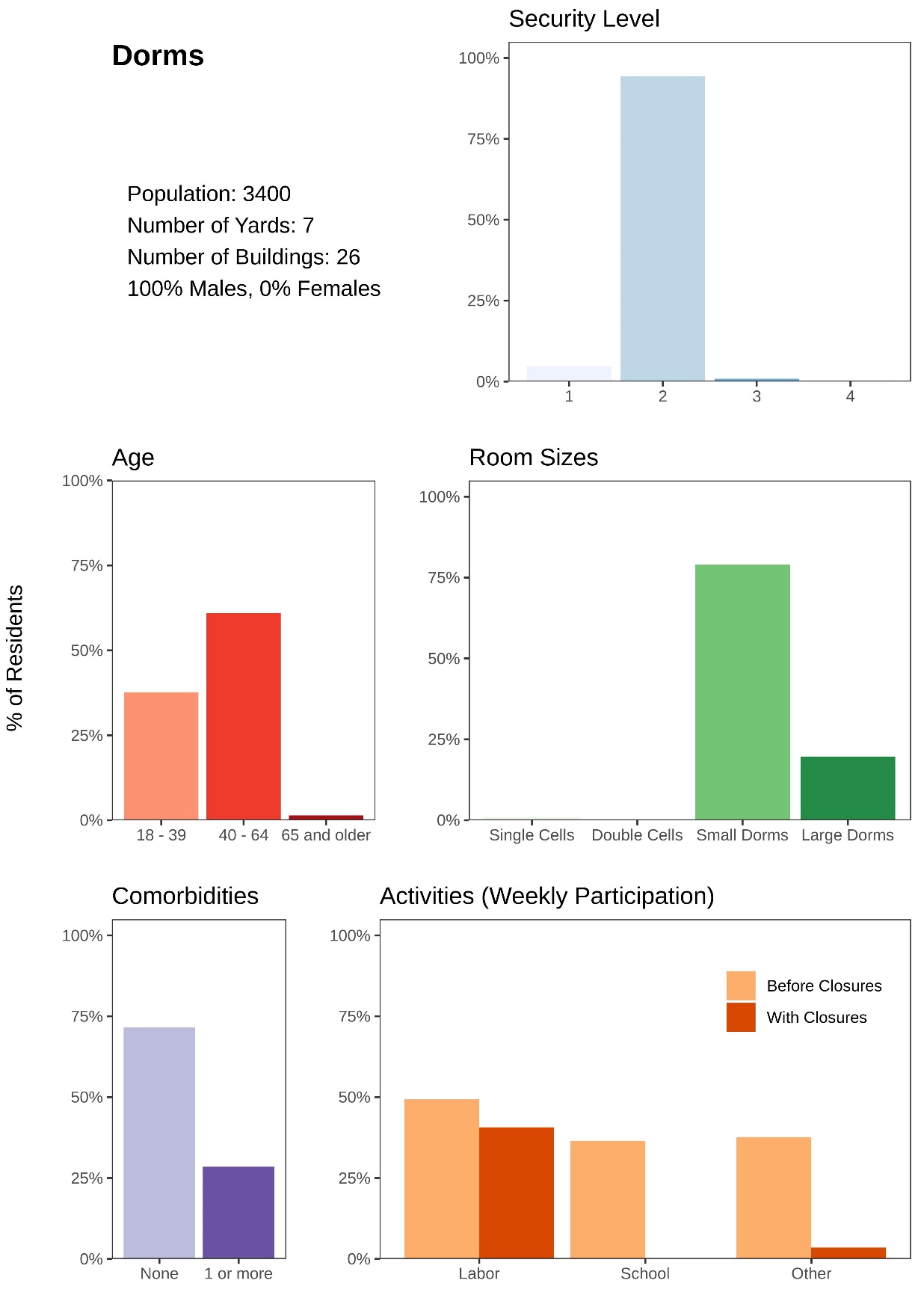

**Figure S7: Medium/mixed security prison with a mix of cells and dorms**

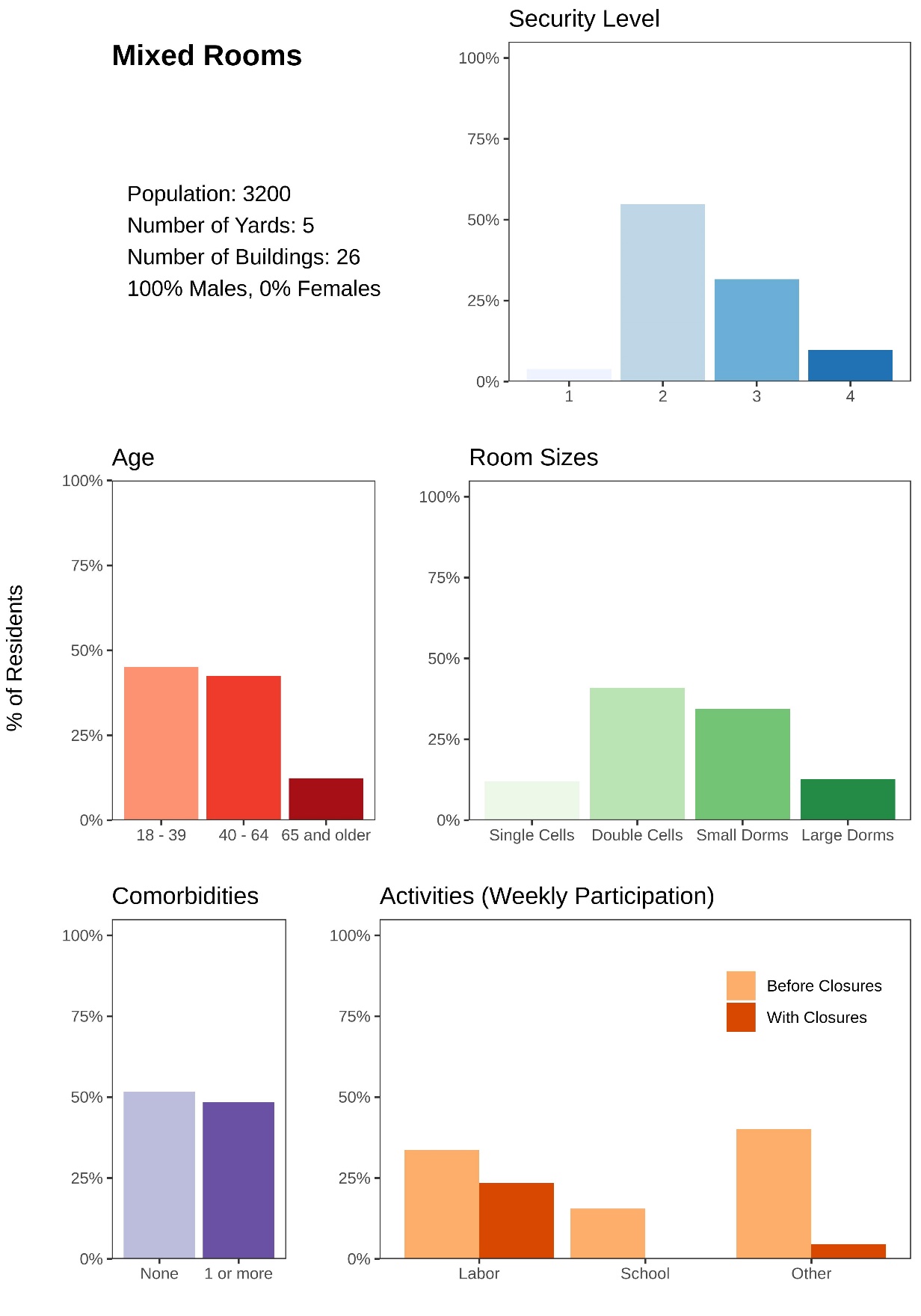

**Figure S8: Higher security prison with mostly cells**

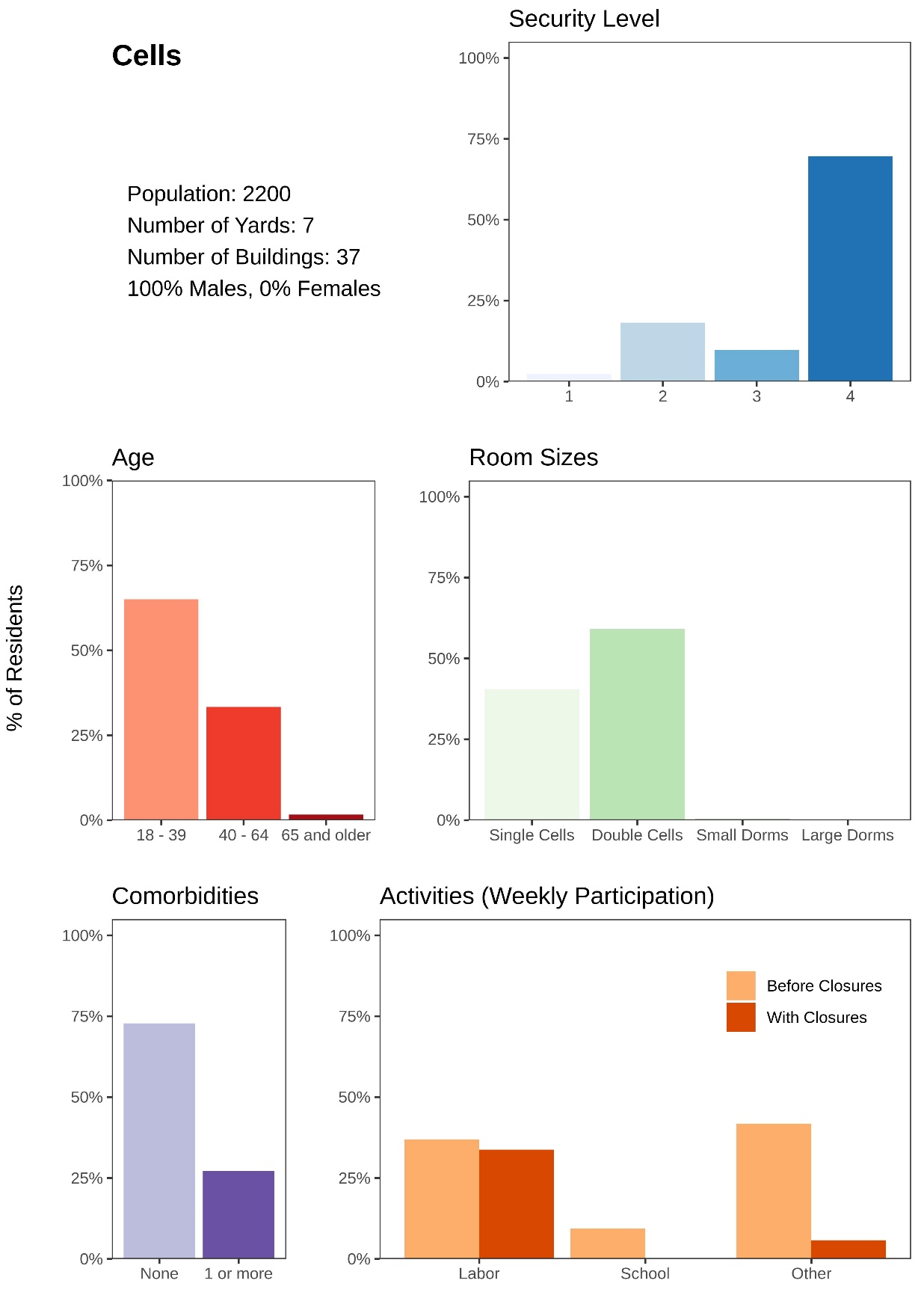

**Figure S9: Women’s prison with mixed security levels and mixed housing**

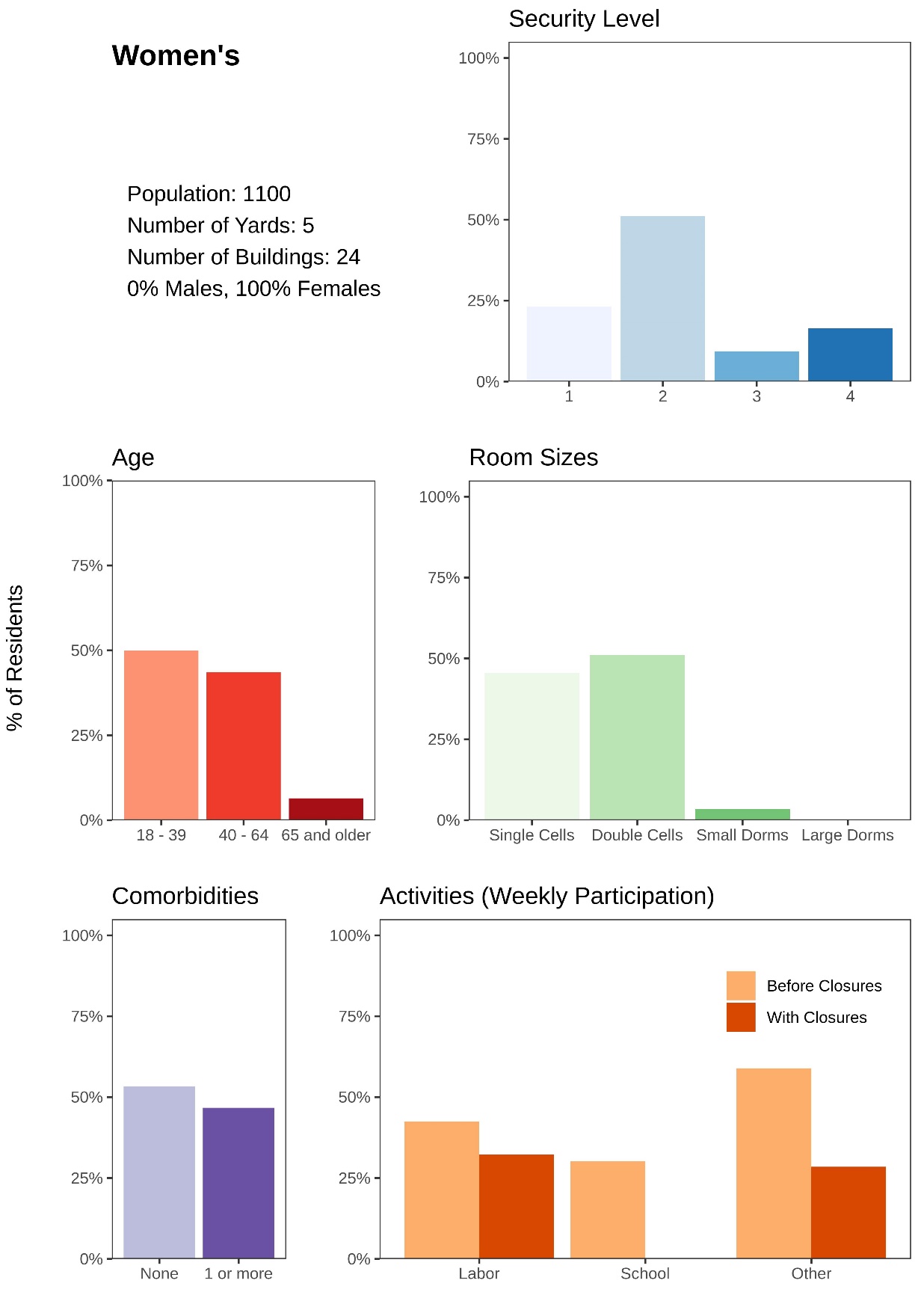

**Figure S10: Medical prison with older and more medically vulnerable residents**

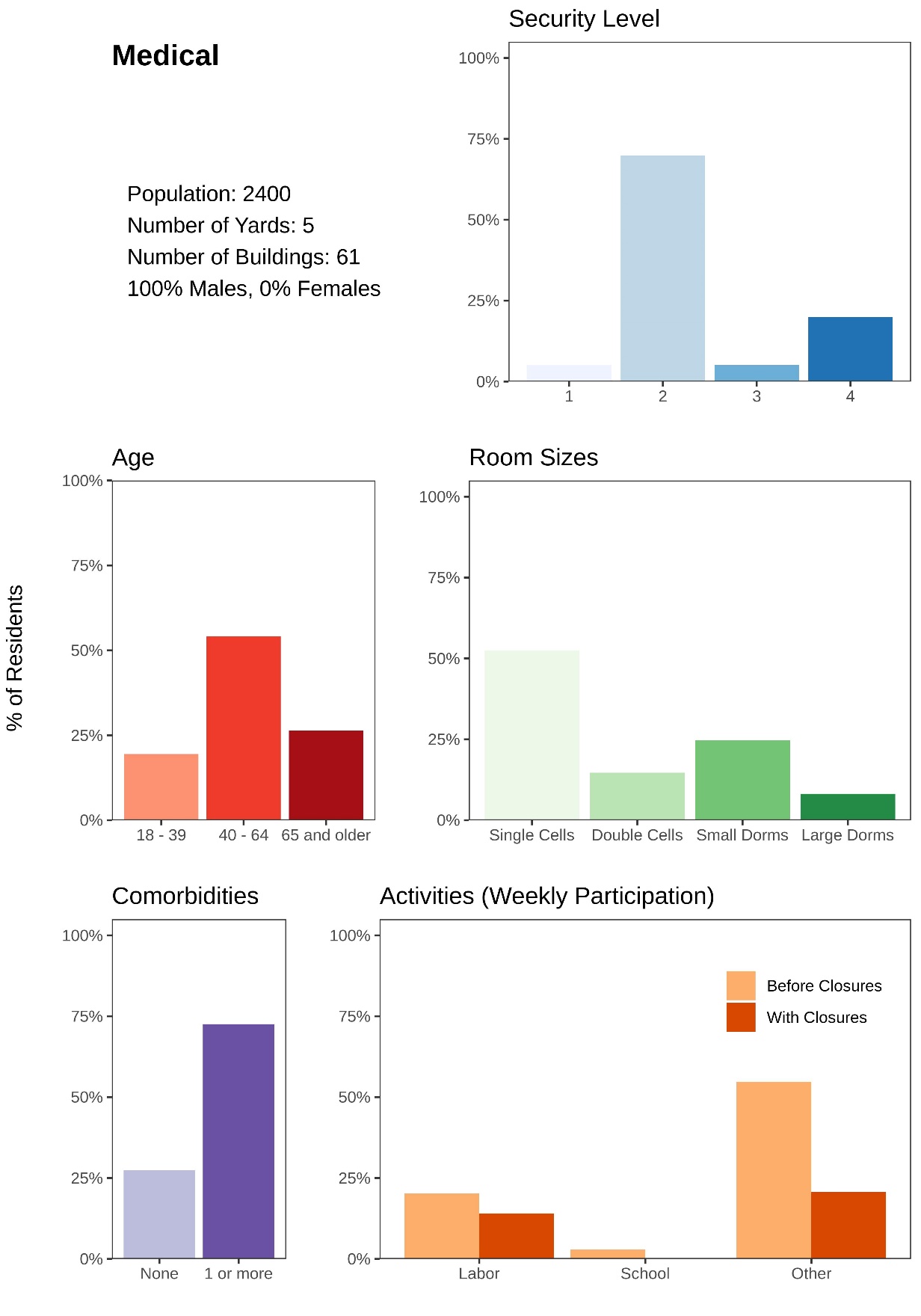

**Model Parameters**

This section describes several model parameters in more detail, including test sensitivity, hospitalization rates, and mortality rates.

*Test Sensitivity*

Test sensitivity by day of infection, based on (7), is described in Table S3.

**Table S3: Probability of testing negative by day of infection (false negative rate)**

| **Day** | **Median [range]** |
| --- | --- |
| 1 | 100% [100-100%] |
| 2 | 100% [95.5-100%] |
| 3 | 97.6% [56.9-99.9%] |
| 4 | 70.4% [29.1-94.0%] |
| 5 | 38.3% [18.3-63.6%] |
| 6 | 24.5% [14.1-39.4%] |
| 7 | 19.8% [12.5-30.3%] |
| 8 | 18.8% [12.1-28.4%] |
| 9 | 19.7% [12.9-29.3%] |
| 10 | 21.8% [14.6-31.8%] |
| 11 | 24.7% [16.9-35.3%] |
| 12 | 28.2% [19.7-39.4%] |
| 13 | 32.3% [23.0-43.8%] |
| 14 | 36.5% [26.5-48.4%] |
| 15 | 40.9% [30.2-52.7%] |
| 16 | 45.2% [34.0-57.0%] |
| 17 | 49.3% [37.9-61.0%] |
| 18 | 53.2% [41.4-64.6%] |
| 19 | 56.7% [45.1-67.8%] |
| 20 | 59.9% [48.5-70.8%] |
| 21 | 62.8% [51.5-73.3%] |

*Resident vaccine acceptance levels*

We model two vaccine coverage scenarios (Table S4). Realistic vaccine acceptance is based on empirical acceptance rates observed among residents of CDCR prisons (8). We calculated the probability of acceptance by age, and then adjusted all the age-based probabilities down by 15 percentage points. This adjustment was made because CDCR initially prioritized highest risk populations and acceptance rates tended to decrease over time as offers were expanded to higher fractions of their total population.

**Table S4: Resident vaccine acceptance by scenario**

| **Resident Age (years)** | **Realistic Vaccine Acceptance** | **Idealized Vaccine Acceptance** |
| --- | --- | --- |
| 18-29 | 36% | 90% |
| 30-39 | 46% | 90% |
| 40-49 | 57% | 90% |
| 50-59 | 66% | 90% |
| 60-69 | 71% | 90% |
| 70-79 | 76% | 90% |
| ≥ 80 | 76% | 90% |

*Resident Covid-19 hospitalization and mortality rates*

For residents, the probability of requiring hospitalization (conditional on having symptomatic Covid-19 infection) and the probability of dying from Covid-19 (conditional on requiring hospitalization) were based on residents’ age and comorbidities. We obtained hospitalization and mortality rates by age from published sources (9,10). We also calculated empirical hospitalization and mortality rates by age and Covid-19 risk score from CDCR data (Figure S11). As mentioned above, the Covid-19 risk score is a metric developed by CDCR to measure the presence of demographic and clinical characteristics identified in the scientific literature as risk factors for severe Covid-19-related illness (Table S2). To adjust the published estimates to reflect prison-specific hospitalization and mortality rates and allow risks of severe outcomes to depend not just on age but also comorbidities, we applied Covid-19 risk score-based multipliers to the published age-based hospitalization and mortality rates. This yielded hospitalization and mortality rates by both age and Covid-19 risk score that were consistent with empirical estimates from CDCR data. The resulting Covid-19 hospitalization and mortality rates are shown in Table S5. Note that hospitalization is modeled as any case severe enough to require hospitalization and is not meant to reflect case detection or hospital capacity.

**Figure S11: Covid-19 hospitalization and mortality among CDCR residents by age and Covid-19 risk score**

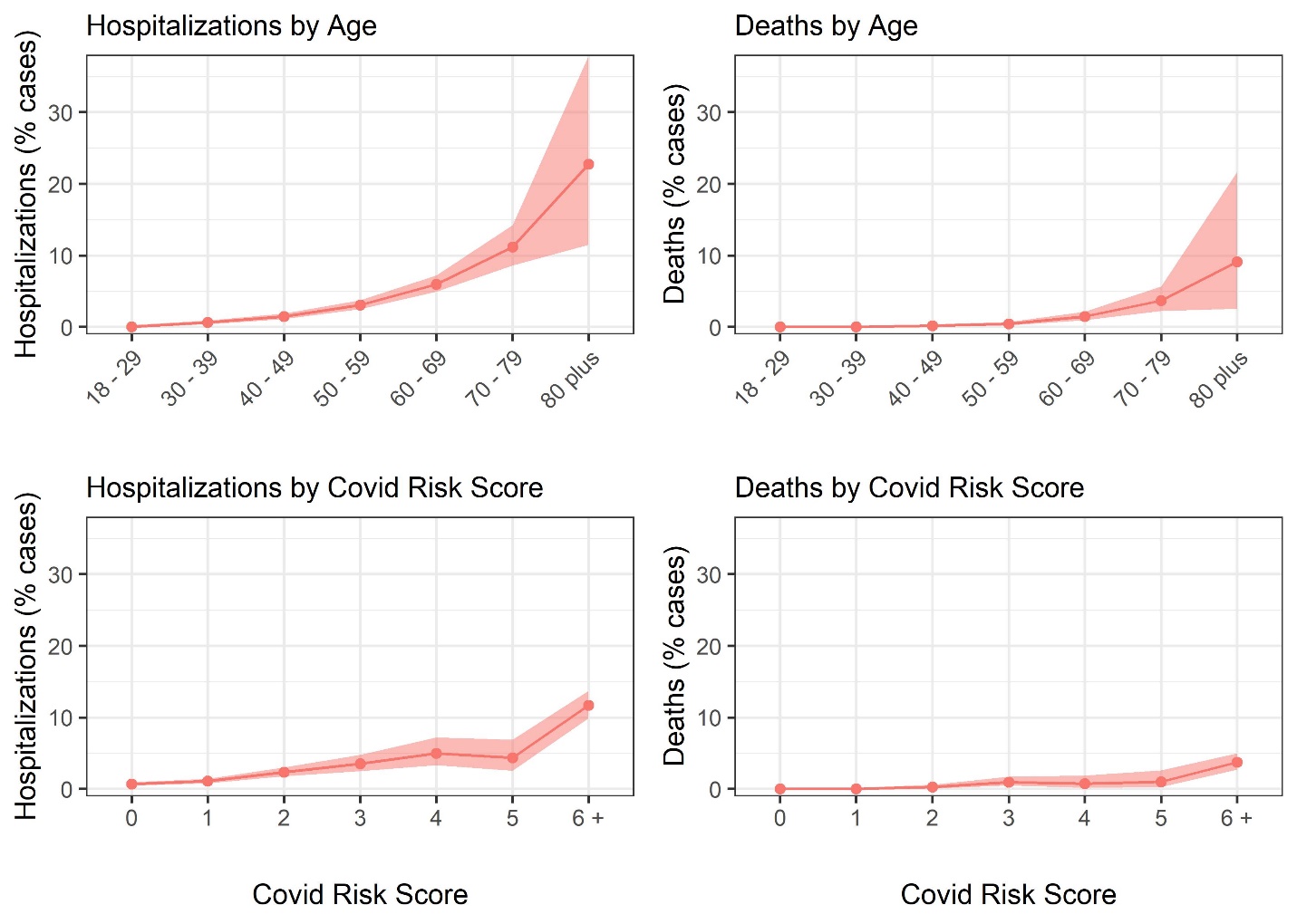

**Table S5: Resident Covid-19 hospitalization and mortality rates**

| **Resident Age (years)** | **Resident Covid-19 Risk Score** | **Probability of requiring hospitalization, conditional on symptomatic infection** | **Probability of death, conditional on requiring hospitalization** |
| --- | --- | --- | --- |
| 18-29 | 0-2 | 0.4% | 3.7% |
|  | 3-7 | 0.8% |  |
|  | ≥ 8 | 1.1% |  |
| 30-39 | 0-2 | 1.0% | 3.8% |
|  | 3-7 | 2.0% |  |
|  | ≥ 8 | 2.9% |  |
| 40-49 | 0-2 | 1.7% | 5.0% |
|  | 3-7 | 3.4% |  |
|  | ≥ 8 | 5.1% |  |
| 50-59 | 0-2 | 3.3% | 9.3% |
|  | 3-7 | 6.5% |  |
|  | ≥ 8 | 9.8% |  |
| 60-69 | 0-2 | 5.5% | 19.7% |
|  | 3-7 | 11.0% |  |
|  | ≥ 8 | 16.4% |  |
| 70-79 | 0-2 | 8.1% | 31.3% |
|  | 3-7 | 16.2% |  |
|  | ≥ 8 | 24.2% |  |
| ≥ 80 | 0-2 | 9.0% | 52.3% |
|  | 3-7 | 18.0% |  |
|  | ≥ 8 | 27.0% |  |

Table shows hospitalization and death probabilities with wild type infection. We model a 1.63 relative increase in the risk of hospitalization and a 1.56 relative increase in the risk of death with variant infection (11).

*Resident background mortality rates*

We estimated background mortality among residents by age and sex based on empirical mortality data from CDCR covering residents of California prisons from November 1, 2016, through October 31, 2019. For males, we used unadjusted rates by age calculated from the CDCR data. For females, who make up less than 5% of the resident population of prisons in California, estimates were noisier because there were relatively fewer deaths. We therefore used background mortality rates from CDC life tables (12) (for females, by age), but adjusted these mortality rates using smoothed hazard rate ratios that reflect higher mortality among incarcerated women compared to the general population. The resulting mortality rates used in the model are shown in Figure S12.

**Figure S12: Unadjusted and adjusted CDCR background mortality rates and CDC background mortality rates**

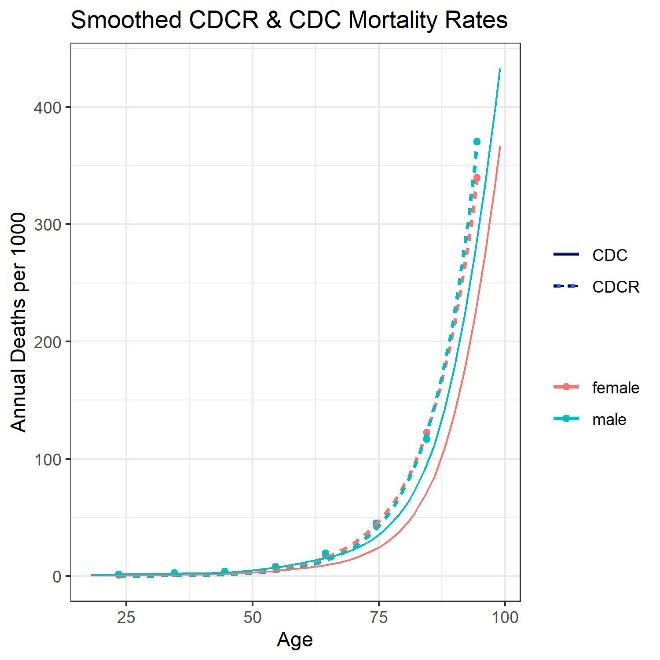

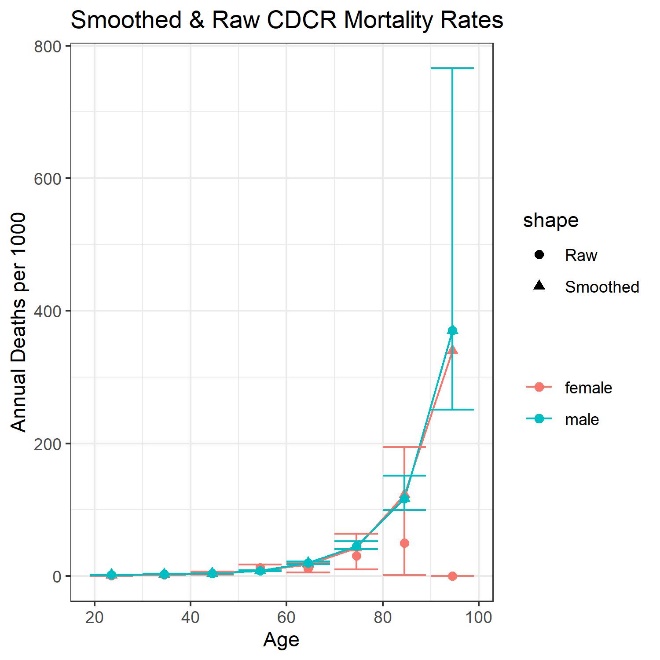

Left panel shows adjusted background mortality rates used in the model (dashed lines) compared to CDC background mortality rates by age and sex. Right panel shows adjusted background mortality rates used in the model (triangles) compared to the raw unadjusted mortality rates (circles).

*Correctional staff Covid-19 population age distribution and mortality rates*

The age and sex distributions of correctional staff were based on census data for correctional workers from the American Community Survey (13). We assumed that there are 6.1 residents per 1 correctional staff, based on data from the Department of Justice which determined the size of the correctional staff for each prison we modeled (14). Background mortality rates were consistent with age-sex-specific background mortality for the US population, obtained from CDC life tables (12).

The probability of dying from Covid-19 (conditional on having symptomatic Covid-19 infection) was based on staff members’ age only and was obtained from public sources (9,10) (Table S5). Hospitalization for staff was not modeled.

**Table S6: Correctional staff parameters**

| **Staff Age (years)** | **Percentage of staff population (total, by age)** | **Percentage of age group that are males** | **Annual background mortality rate** | **Probability of death, conditional on symptomatic wild type Covid-19 infection** |
| --- | --- | --- | --- | --- |
| 18-29 | 10% | 76.6% | 0.0012 | 0.03% |
| 30-39 | 15% | 79.7% | 0.0019 | 0.08% |
| 40-49 | 25% | 77.9% | 0.0030 | 0.17% |
| 50-59 | 25% | 79.3% | 0.0070 | 0.64% |
| 60-69 | 25% | 80.8% | 0.0145 | 2.37% |

We model a 1.56 relative increase in the risk of death with variant infection (11).

**Supplemental References**

1.	Keeling MJ, Rohani P. Modeling Infectious Diseases in Humans and Animals [Internet]. Princeton University Press; 2007 [cited 2021 Mar 19]. Available from: https://press.princeton.edu/books/hardcover/9780691116174/modeling-infectious-diseases-in-humans-and-animals

2.	Hu H, Nigmatulina K, Eckhoff P. The scaling of contact rates with population density for the infectious disease models. Mathematical Biosciences. 2013 Aug 1;244(2):125–34.

3.	Dalziel BD, Kissler S, Gog JR, Viboud C, Bjørnstad ON, Metcalf CJE, et al. Urbanization and humidity shape the intensity of influenza epidemics in U.S. cities. Science. 2018 Oct 5;362(6410):75–9.

4.	Li Q, Guan X, Wu P, Wang X, Zhou L, Tong Y, et al. Early Transmission Dynamics in Wuhan, China, of Novel Coronavirus-Infected Pneumonia. N Engl J Med. 2020 Mar 26;382(13):1199–207.

5.	Alimohamadi Y, Taghdir M, Sepandi M. Estimate of the Basic Reproduction Number for COVID-19: A Systematic Review and Meta-analysis. J Prev Med Public Health. 2020 May;53(3):151–7.

6.	Chin ET, Ryckman T, Prince L, Leidner D, Alarid-Escudero F, Andrews JR, et al. Covid-19 in the California State Prison System: An Observational Study of Decarceration, Ongoing Risks, and Risk Factors. medRxiv. 2021 Mar 8;2021.03.04.21252942.

7.	Kucirka LM, Lauer SA, Laeyendecker O, Boon D, Lessler J. Variation in False-Negative Rate of Reverse Transcriptase Polymerase Chain Reaction-Based SARS-CoV-2 Tests by Time Since Exposure. Ann Intern Med. 2020 Aug 18;173(4):262–7.

8.	Chin ET, Leidner D, Ryckman T, Liu Y, Prince L, Alarid-Escudero F, et al. Covid-19 Vaccine Acceptance among Residents of California State Prisons. Forthcoming. 2021 Mar;

9.	Verity R, Okell LC, Dorigatti I, Winskill P, Whittaker C, Imai N, et al. Estimates of the severity of coronavirus disease 2019: a model-based analysis. The Lancet Infectious Diseases. 2020 Jun 1;20(6):669–77.

10.	Model Parameters [Internet]. [cited 2021 Apr 1]. Available from: https://mrc-ide.github.io/squire/articles/parameters.html

11.	Tuite AR, Fisman DN, Odutayo A, Bobos P, Allen V, Bogoch II, et al. COVID-19 Hospitalizations, ICU Admissions and Deaths Associated with the New Variants of Concern [Internet]. Ontario COVID-19 Science Advisory Table. [cited 2021 Apr 8]. Available from: https://covid19-sciencetable.ca/sciencebrief/covid-19-hospitalizations-icu-admissions-and-deaths-associated-with-the-new-variants-of-concern/

12.	US Centers for Disease Control and Prevention. Life Tables [Internet]. 2019 [cited 2021 Mar 9]. Available from: https://www.cdc.gov/nchs/products/life_tables.htm

13.	US Census Bureau. American Community Survey Data [Internet]. The United States Census Bureau. [cited 2021 Mar 9]. Available from: https://www.census.gov/programs-surveys/acs/data.html

14.	Staphan J. Census of State and Federal Correctional Facilities, 2005 [Internet]. Bureau of Justice Statistics; Available from: https://www.bjs.gov/content/pub/pdf/csfcf05.pdf

**Supplementary Results**

**Appendix Figure S13: Cumulative resident cases requiring hospitalization per 1000 infections over 200 days by in-person activity status, widespread use on NPIs, and baseline immunity, conditional on introduction of a single new variant infection**

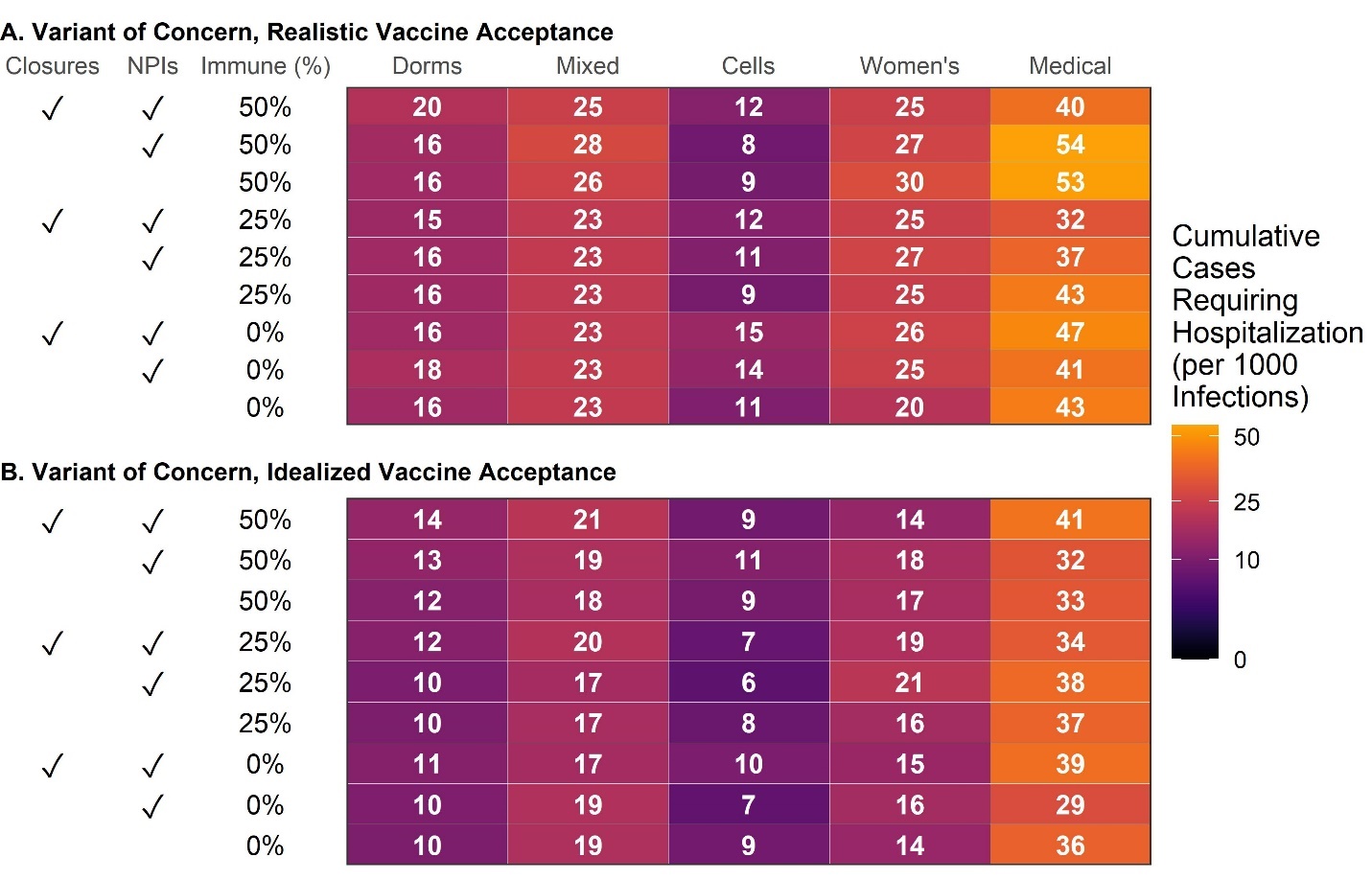

Figure shows average cumulative severe cases (requiring hospitalization) among infected residents (not all residents) across 500 model simulations over 200 days for each scenario shown.

**Appendix Figure S14: Upper bound on cumulative resident cases requiring hospitalization over 200 days by in-person activity status, widespread use on NPIs, and baseline immunity, conditional on introduction of a single variant infection**

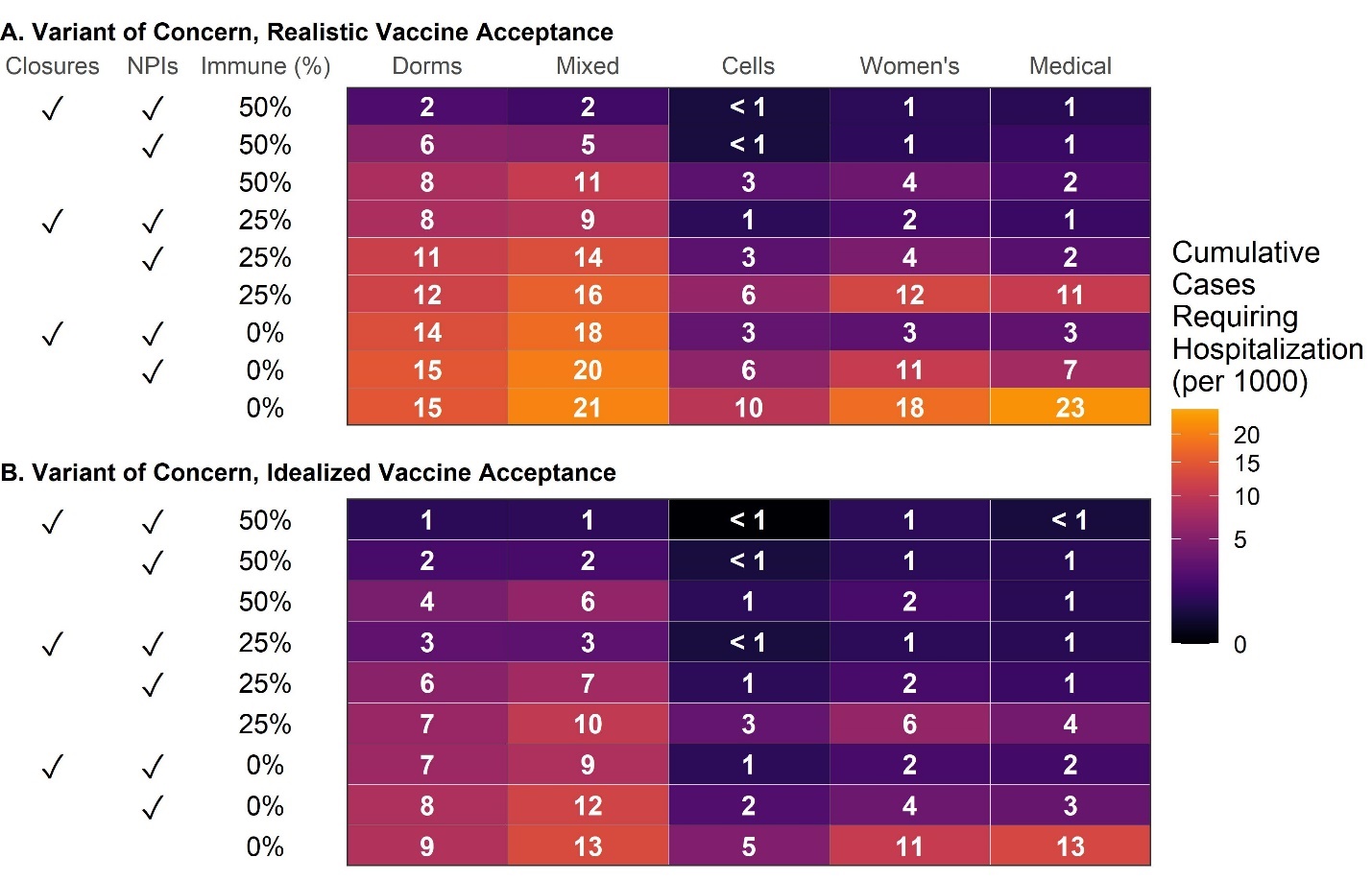

Figure shows the 95^th^ percentile of cumulative severe cases (requiring hospitalization) among residents across 500 model simulations over 200 days for each scenario shown.

**Appendix Figure S15: Median reduction in resident infections over 200 days from vaccination compared to no vaccination, by in-person activity status, widespread use on NPIs, and baseline immunity, conditional on introduction of a single new variant infection**

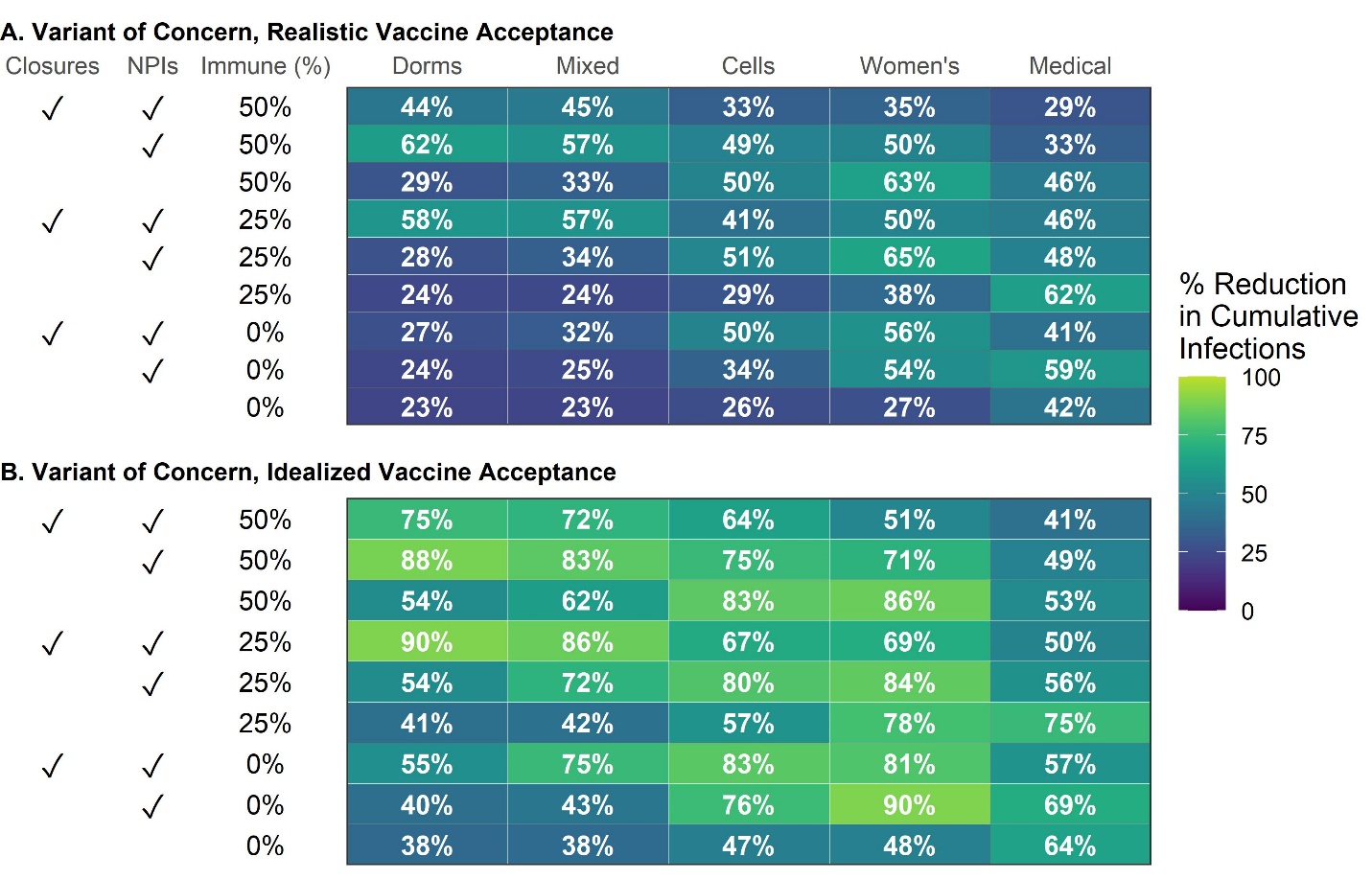

Figure shows the median reduction from vaccination (compared to no vaccination) in cumulative infections among residents across 500 model simulations over 200 days for each scenario shown.

**Appendix Figure S16: Median reduction in resident cases requiring hospitalization over 200 days from vaccination compared to no vaccination, by in-person activity status, widespread use on NPIs, and baseline immunity, conditional on introduction of a single new variant infection**

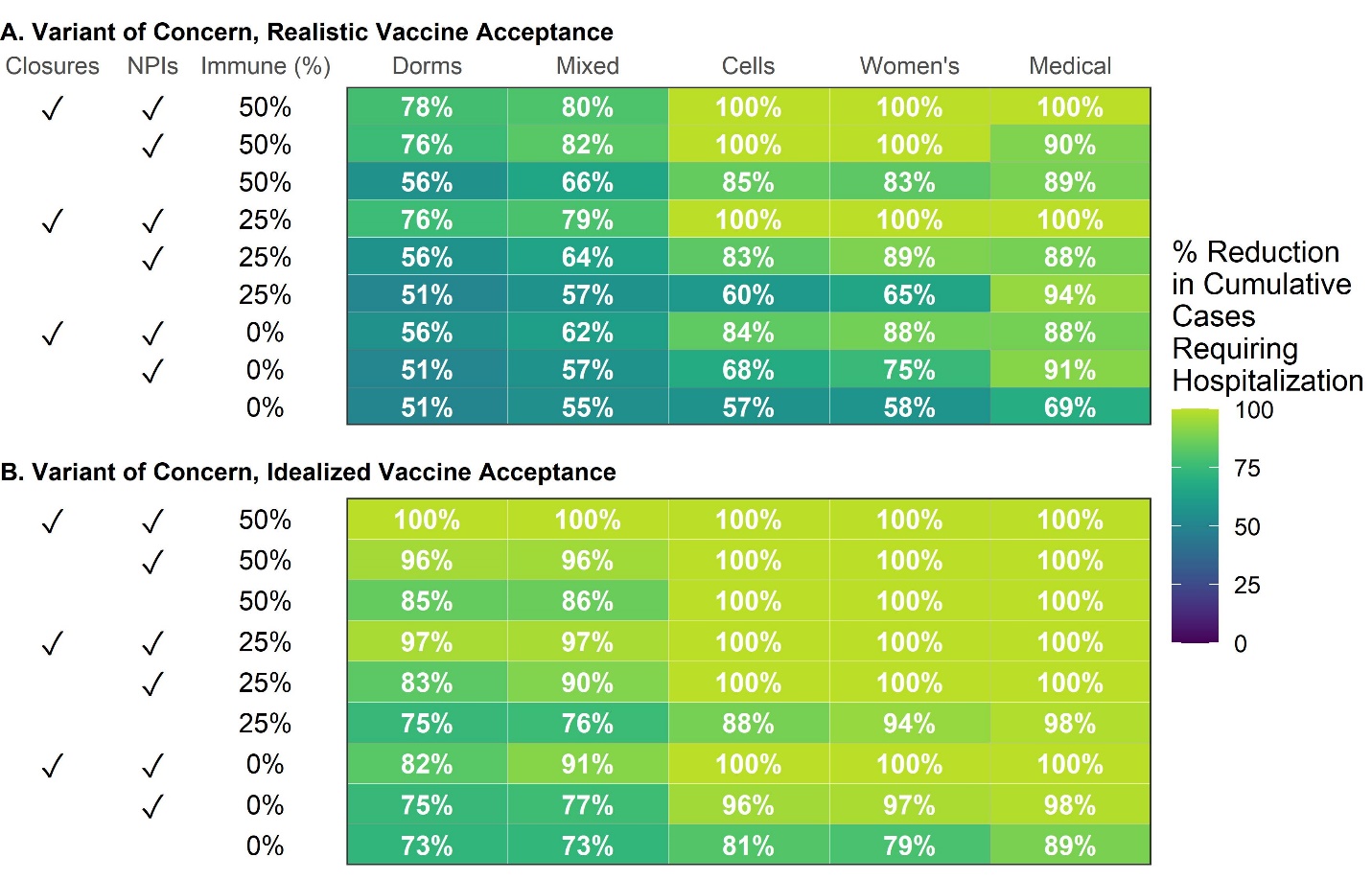

Figure shows the median reduction from vaccination (compared to no vaccination) in cumulative severe cases (requiring hospitalization) among residents across 500 model simulations over 200 days for each scenario shown.

**Appendix Figure S17: Cumulative resident infections over 200 days by in-person activity status, widespread use on NPIs, and baseline immunity, conditional on introduction of a single wild type infection**

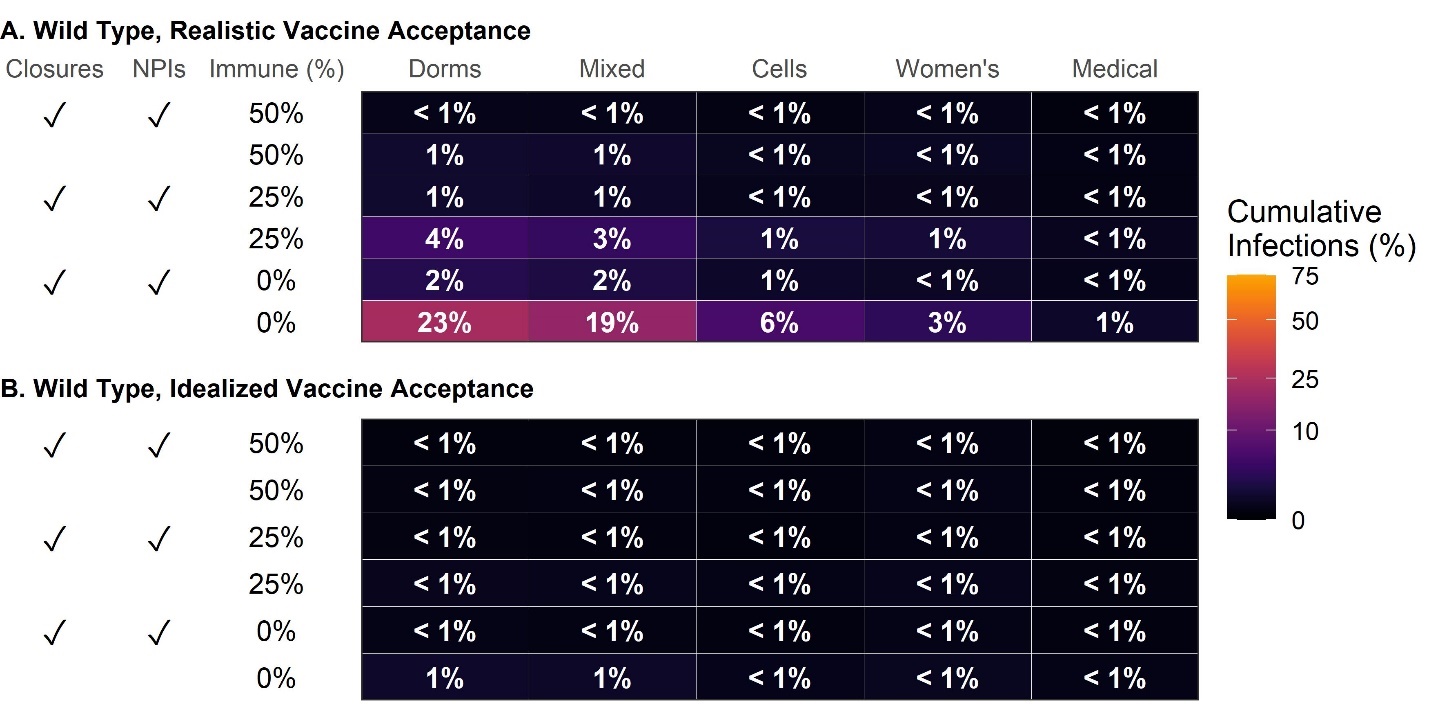

Figure shows average cumulative infections among residents across 500 model simulations over 200 days for each scenario shown.

**Appendix Figure S18: Cumulative resident cases requiring hospitalization over 200 days by in-person activity status, widespread use on NPIs, and baseline immunity, conditional on introduction of a single wild type infection**

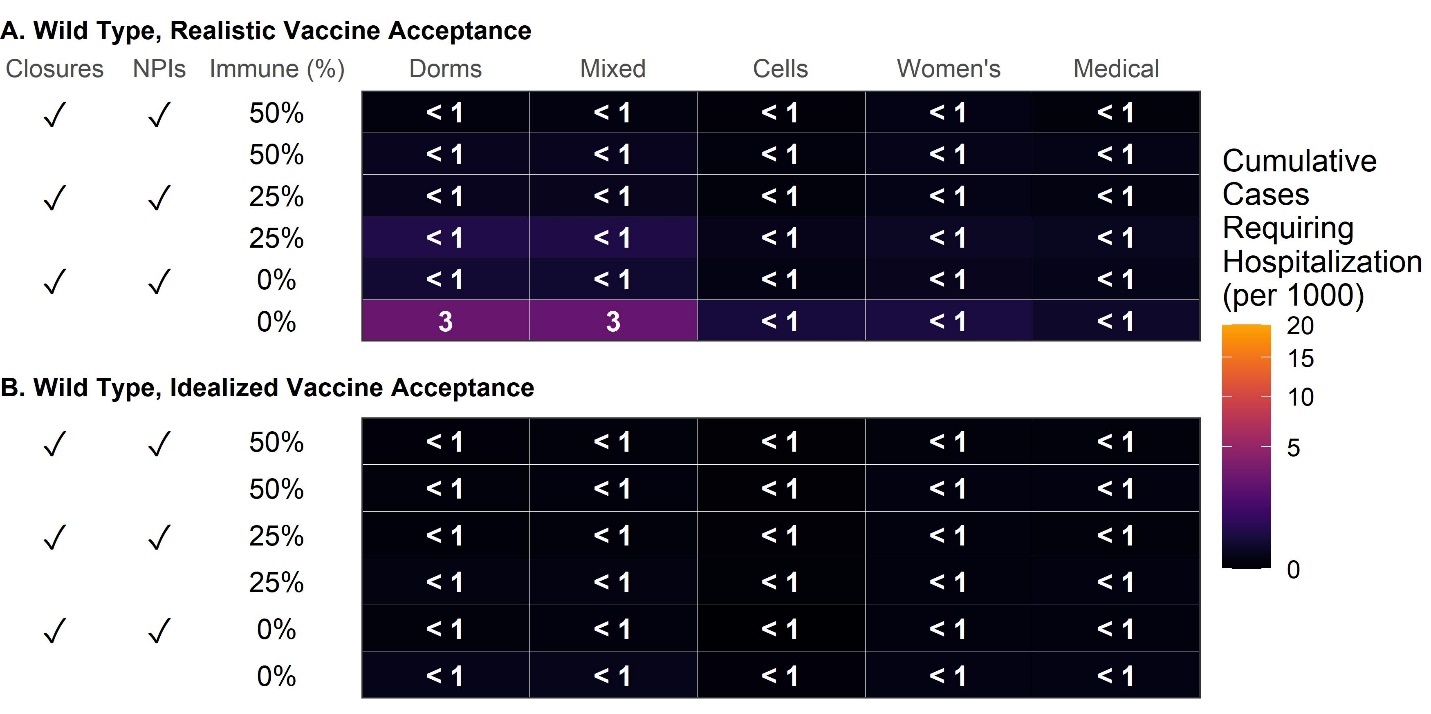

Figure shows average cumulative severe cases (requiring hospitalization) among residents across 500 model simulations over 200 days for each scenario shown.

**Appendix Figure S19: Cumulative resident infections over 200 days by in-person activity status, widespread use on NPIs, and baseline immunity, conditional on continual importation of variant infections**

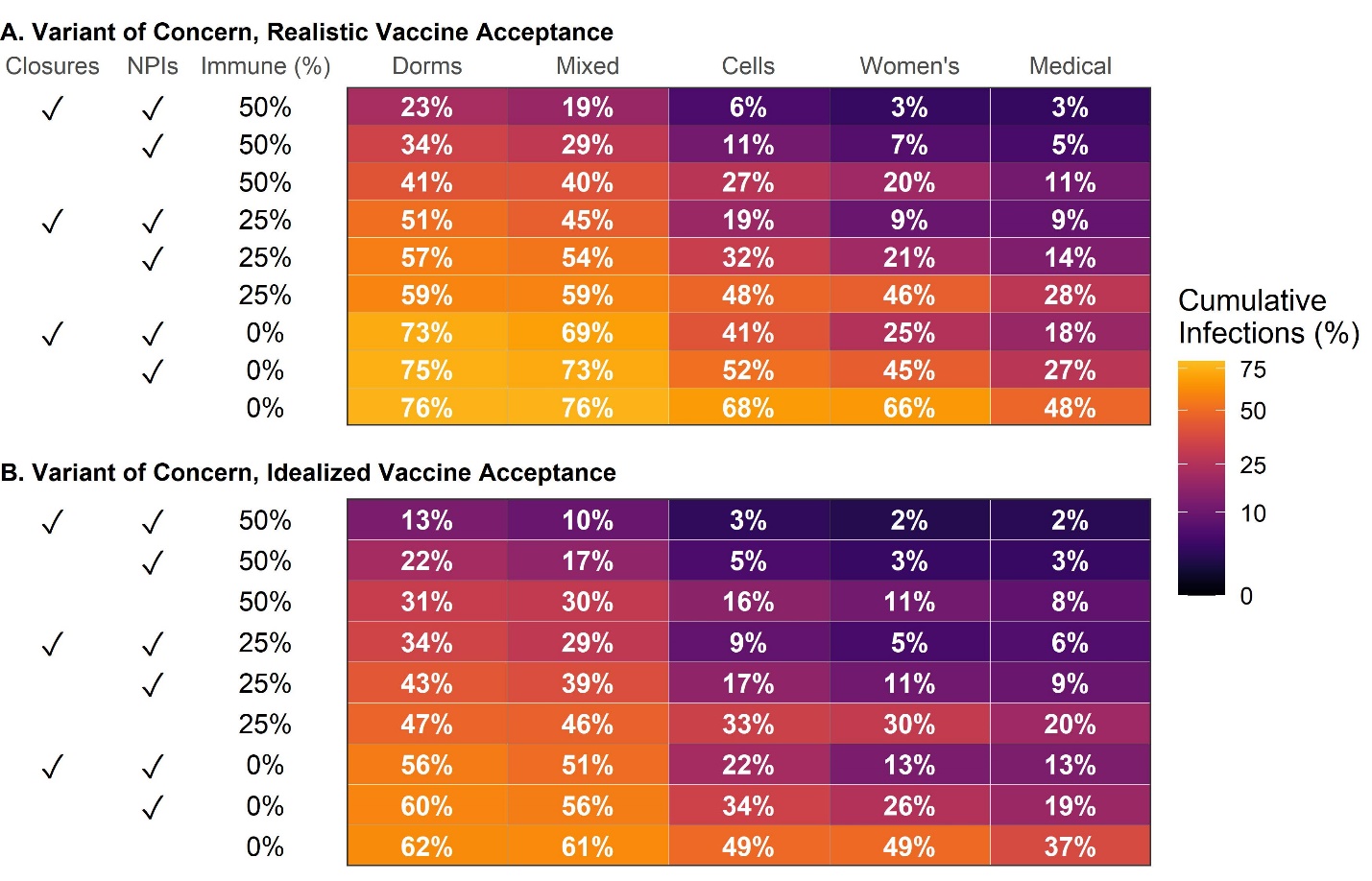

Figure shows average cumulative infections among residents across 500 model simulations over 200 days with 0.1% daily incidence among susceptible staff members for each scenario shown.

**Appendix Figure S20: Cumulative resident cases requiring hospitalization over 200 days by in-person activity status, widespread use on NPIs, and baseline immunity, conditional on continual importation of variant infections**

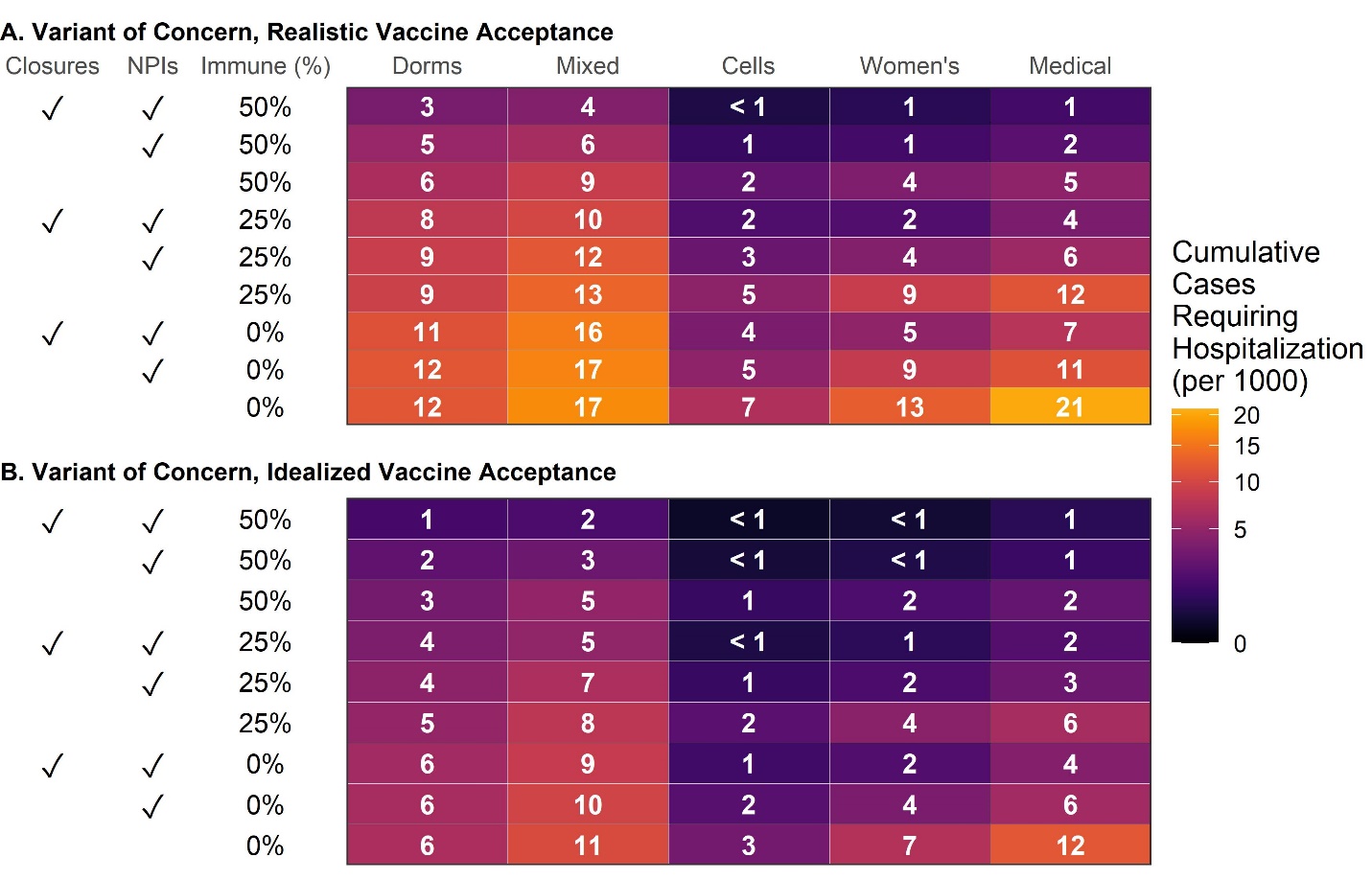

Figure shows average cumulative severe cases (requiring hospitalization) among residents across 500 model simulations over 200 days with 0.1% daily incidence among susceptible staff members for each scenario shown.

**Appendix Figure S21: Cumulative resident infections over 200 days by in-person activity status, widespread use on NPIs, and baseline immunity, conditional on introduction of a single new variant infection concurrent with vaccination scale-up**

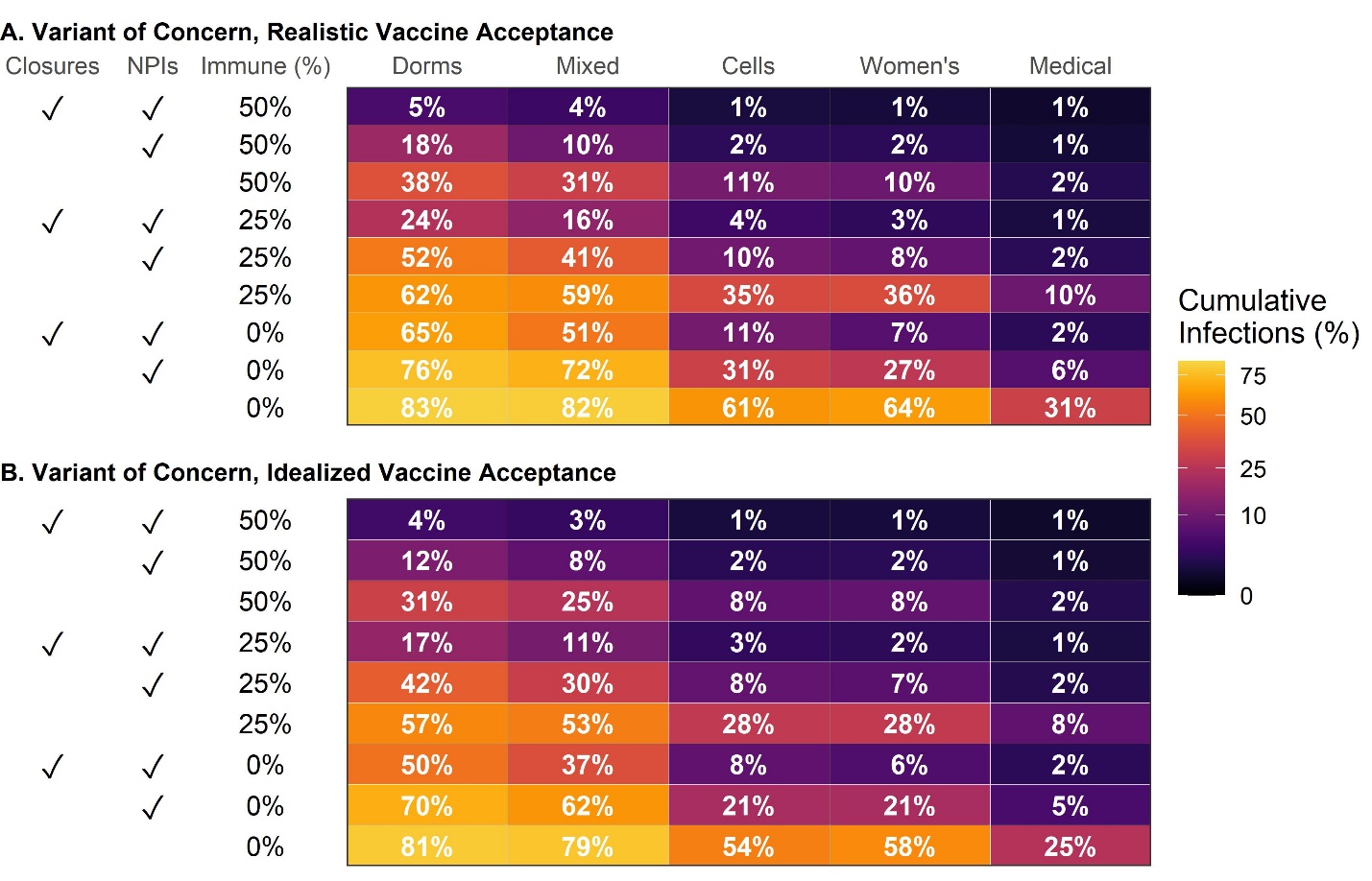

Figure shows average cumulative infections among residents across 500 model simulations over 200 days for each scenario shown. Vaccination of residents is modeled as beginning on the same day that an infection is introduced to the prison (in the main analysis, vaccination is scaled-up prior to introduction).

**Appendix Figure S22: Cumulative resident cases requiring hospitalization over 200 days by in-person activity status, widespread use on NPIs, and baseline immunity, conditional on introduction of a single new variant infection concurrent with vaccination scale-up**

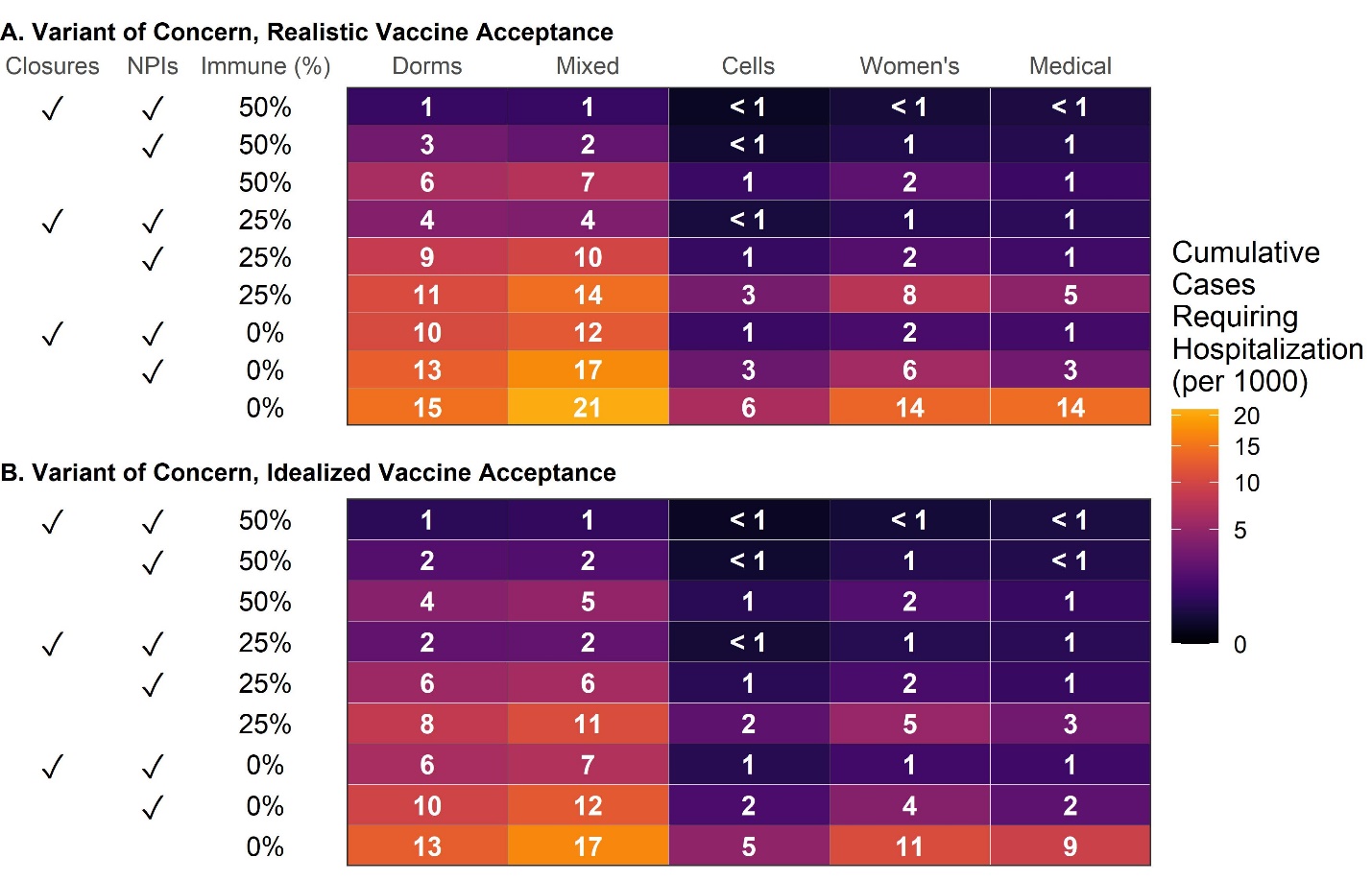

Figure shows average cumulative severe cases (requiring hospitalization) among residents across 500 model simulations over 200 days for each scenario shown. Vaccination of residents is modeled as beginning on the same day that an infection is introduced to the prison (in the main analysis, vaccination is scaled-up prior to introduction).

**Appendix Figure S23: Cumulative resident infections over 200 days by in-person activity status, widespread use on NPIs, and baseline immunity, conditional on introduction of a single new variant infection and 80% staff vaccination coverage**

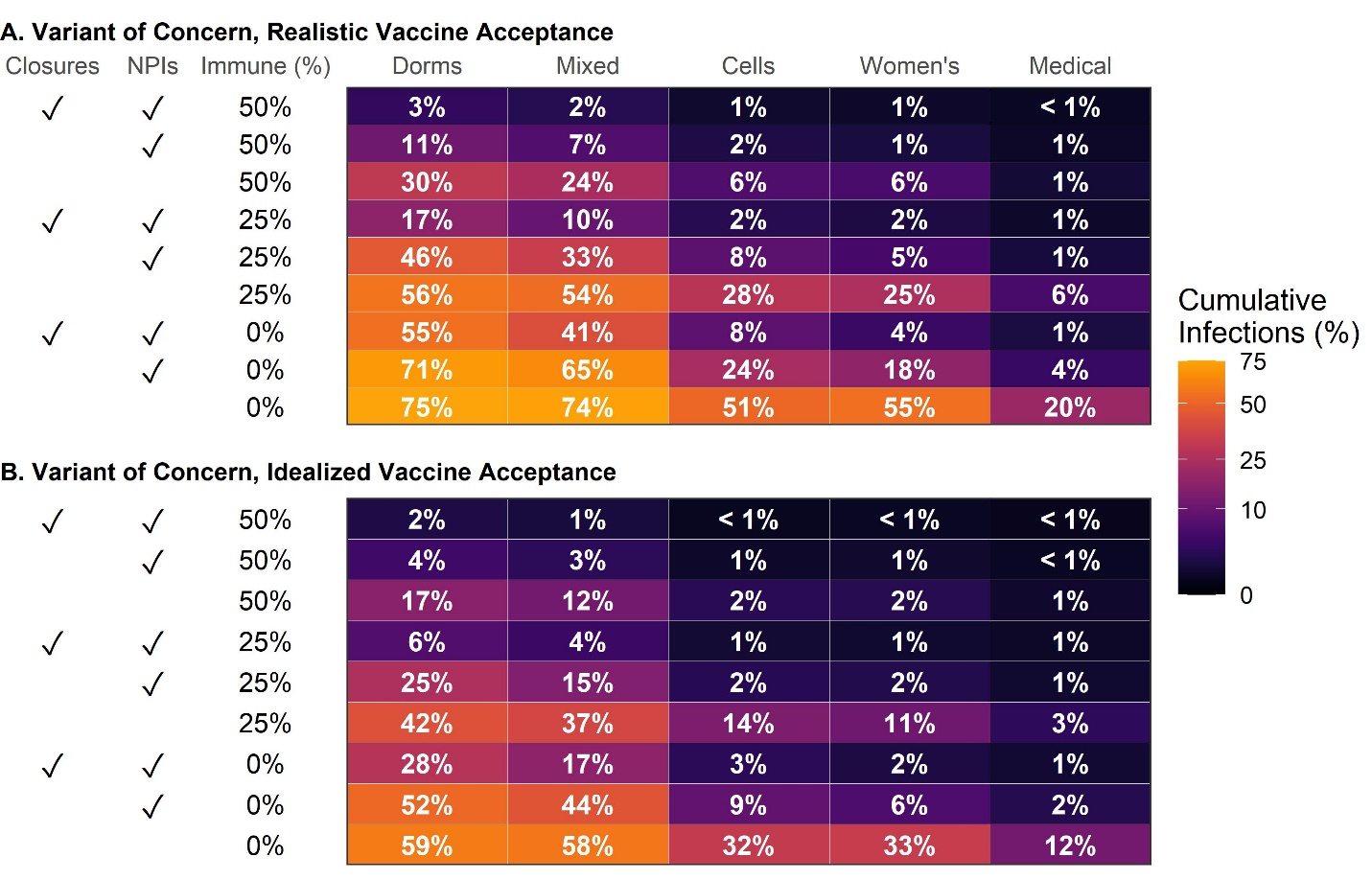

Figure shows average cumulative infections among residents across 500 model simulations over 200 days for each scenario shown. Staff vaccination coverage is set at 80% (compared to 40% in the main analysis).

**Appendix Figure S24: Cumulative resident cases requiring hospitalization over 200 days by in-person activity status, widespread use on NPIs, and baseline immunity, conditional on introduction of a single new variant infection and 80% staff vaccination coverage**

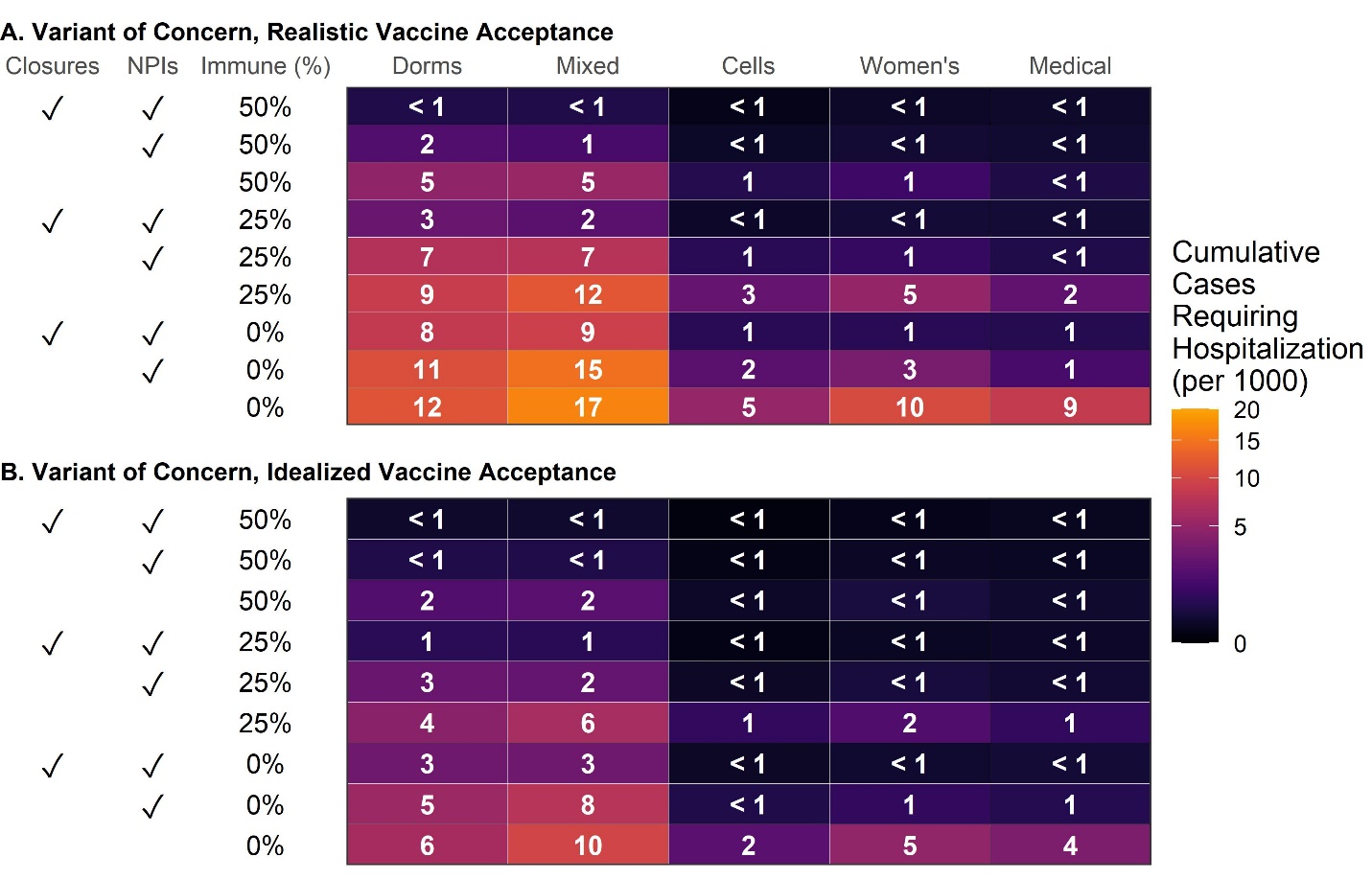

Figure shows average cumulative severe cases (requiring hospitalization) among residents across 500 model simulations over 200 days for each scenario shown. Staff vaccination coverage is set at 80% (compared to 40% in the main analysis).
